# Sex-aware Cross-tissue Regulatory Transformer Identified Sexually Dimorphic Alzheimer’s Disease Risk Loci and Causal Cellular Circuit

**DOI:** 10.64898/2026.09.08.26362488

**Authors:** Zhi’ang Cheng, Seno Judaprawira, James Perez, Adam Goldstein

## Abstract

Sex differences in Alzheimer’s disease susceptibility remain poorly understood at the level of gene regulation. Here we analyzed 449,335 individuals of European ancestry and identified 24 variants with genotype-by-sex interactions at genome-wide significance. We developed STAGE-AD, a sex-conditioned Transformer integrating association statistics with cross-tissue molecular quantitative trait loci, functional annotations, epigenomic profiles, single-cell annotations and neuropathology characteristics. Statistical-genetic analyses in independent cohorts supported 236 genes at 106 prioritized loci. Female-biased loci implicated lipid-responsive microglia and reactive astrocytes, whereas male-biased loci implicated vascular endothelium and oligodendrocytes. Corresponding genetic modules were associated with tau pathology and episodic memory decline, and with white matter injury and executive decline, respectively. Integration of molecular and neuropathological evidence prioritized 11 candidate regulatory chains linking variants to cellular contexts and disease phenotypes, which were validated in NPAD cohort. These findings delineate sex-differentiated regulatory contexts within a largely shared genetic architecture of Alzheimer’s disease and nominate mechanisms for experimental investigation.

## Introduction

Alzheimer’s disease (AD) is the most common cause of dementia in older adults and is characterized by amyloid-β deposition, tau pathology, synaptic dysfunction, neuroinflammation and neurodegeneration. Large-scale genetic studies have identified AD risk loci implicating lipid metabolism, amyloid and tau biology, innate immunity, endocytosis, microglial function and vascular pathways^1,2^. A recent consensus meta-analysis of genome-wide association studies (GWAS) further expanded the genetic architecture of AD and related dementias^3^. These findings support a highly polygenic architecture, but the regulatory and cellular processes through which genetic variation influences AD susceptibility remain incompletely resolved.

Most AD-associated variants map to non-coding regions, making it difficult to infer causal variants, target genes and effector tissues from genomic proximity alone. Statistical fine-mapping, colocalization^4^, transcriptome-wide association studies (TWAS)^5^ and pathway enrichment analyses can prioritize candidate variants, genes and biological processes. These approaches are often applied as separate analytical layers, leaving the links among variants, regulatory elements, target genes, tissues, cell types and disease-relevant cellular states fragmented. Consequently, many AD loci retain multiple plausible effector genes or context-dependent regulatory mechanisms, particularly in regions of complex linkage disequilibrium (LD).

Sex is an important but incompletely resolved source of heterogeneity in AD. AD prevalence is higher among women than men^6^, although differences in survival account for part of this disparity, and the associations of APOE ε4 with AD biomarkers and tau pathology may differ between females and males^7^. Most large-scale AD GWAS have treated sex primarily as a covariate. Recent sex-stratified, genotype-by-sex interaction and X-chromosome-wide analyses have begun to characterize shared and sex-differentiated genetic effects^8–10^, but association-level analyses do not identify the regulatory elements, tissues or cell types through which these effects may operate. Connecting sex-aware genetic associations to candidate regulatory and cellular contexts therefore remains a central challenge.

Deep learning provides one approach to modelling dependencies across heterogeneous genomic data. Sequence-based models such as Enformer and Borzoi predict functional genomic readouts from DNA sequence, whereas single-cell foundation models such as scGPT learn representations from large-scale cellular datasets^11–13^. Deep-learning approaches may also improve GWAS-based variant prioritization by incorporating biological annotations^14^. However, existing models were developed primarily for sequence-to-function prediction or cellular representation learning and do not jointly integrate AD GWAS, sex-stratified association statistics, fine-mapped variants, multi-tissue quantitative trait loci (QTLs), regulatory-element maps and AD-relevant single-cell states. To our knowledge, no established framework explicitly treats sex as a conditioning regulatory context when prioritizing candidate variant-to-gene mechanisms across tissues and cell types in AD.

To address this gap, we developed STAGE-AD, a **S**ex-aware **T**issue-regulatory **A**ttention model for **GE**netic discovery in **AD** (**Fig. 1a**, **Supplementary Fig. S1**). The model integrates sex-stratified AD GWAS and genotype-by-sex (G×S) interaction statistics, multi-tissue QTLs, enhancer–gene maps, functional annotations, AD single-cell transcriptomic and epigenomic profiles, biomarker and neuropathology outcomes. It represents variants, regulatory elements, genes, tissues, cell types and sex contexts as structured sequences and uses self-attention and sex-conditioned cross-attention to prioritize shared, female-biased, male-biased and G×S-interaction mechanisms. We evaluated these priorities using complementary evidence from expression- and splicing-based TWAS, cell-type enrichment, cross-tissue colocalization, partitioned heritability and replication analyses in external independent cohorts. We further tested associations between prioritized sex-aware genetic modules and AD neuropathology and cognitive phenotypes. This framework provides a testable, sex-aware map of candidate regulatory mechanisms and will inform future studies investigating more targeted and precise therapeutics to improve AD among sex-specific genetic risk profiles.

**Fig. 1.**
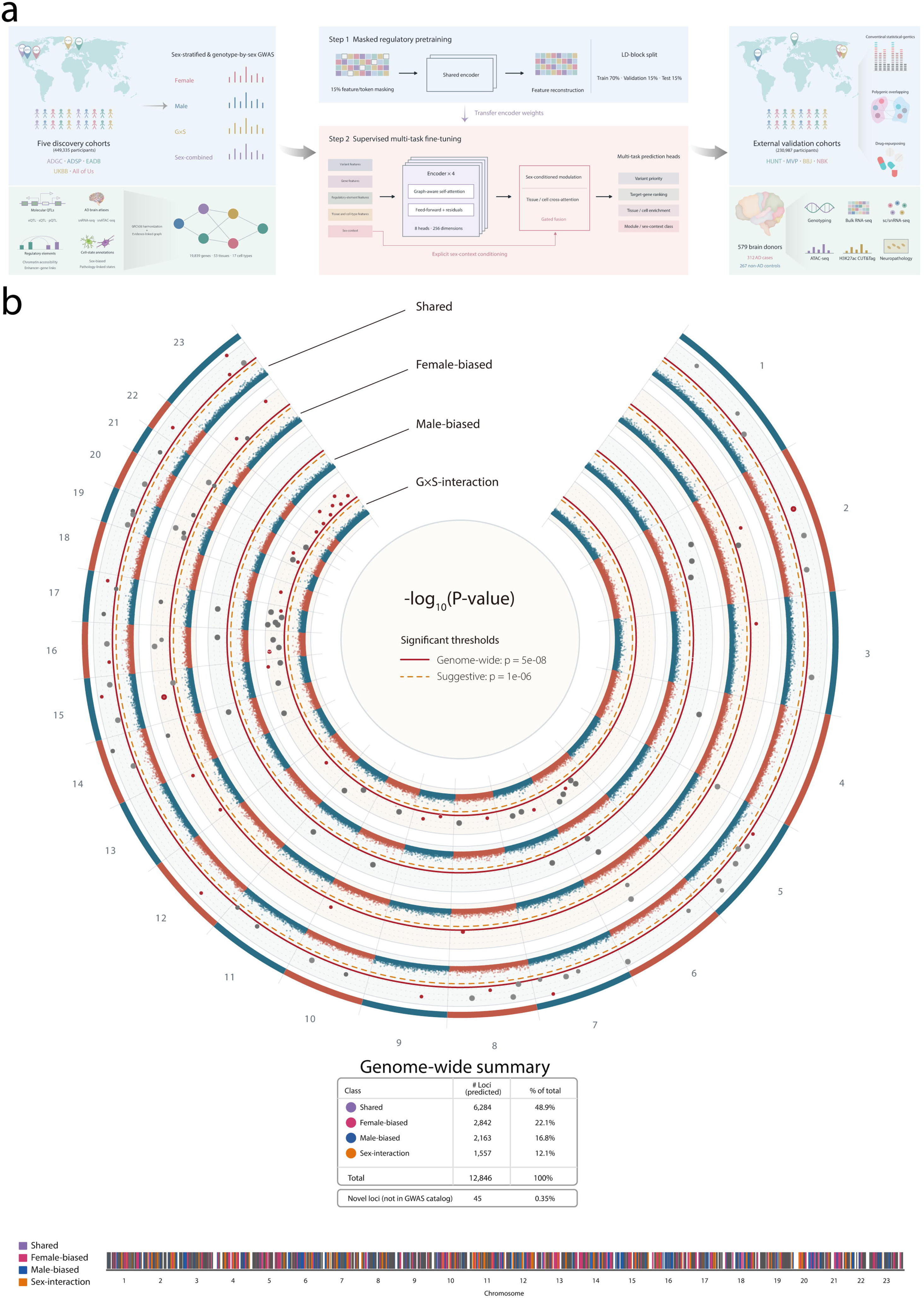
Overview of study design and genome-wide distribution prioritization of Alzheimer’s disease loci by STAGE-AD. **a**, Study design and STAGE-AD workflow. Sex-stratified, sex-combined and genotype-by-sex genome-wide association analyses included 449,335 participants from five discovery cohorts (ADGC, ADSP, EADB, UKBB and All of Us). Association data were integrated with molecular quantitative trait loci, brain single-cell atlases, regulatory annotations and cell-state information. Masked regulatory pretraining was followed by supervised multitask fine-tuning with sex-conditioned, graph-aware attention to prioritize variants, target genes, tissues, cell types and sex-context modules. Linkage disequilibrium blocks were partitioned into training, validation and test sets (70%, 15% and 15%, respectively). External analyses used HUNT, MVP, BBJ and NBK (230,987 participants). The independent NPAD cohort comprised 579 brain donors (312 AD cases and 267 non-AD controls) and provided genetic, transcriptomic, epigenomic and neuropathological evidence. **b**, Circular Manhattan plots of discovery association statistics. From outermost to innermost, tracks show shared, female, male and genotype-by-sex interaction analyses. Each point represents a variant, with radial position indicating −log₁₀(P). Red and dark-grey highlighted points denote variants at loci classified in this study as novel and previously reported, respectively; other points are colored by chromosome. Solid red and dashed orange lines indicate genome-wide significance (P = 5E-08) and suggestive association (P = 1E-06) thresholds, respectively. The inset summarizes model-assigned classes. The lower strip shows their chromosome-ordered distribution: shared (purple), female-biased (magenta), male-biased (blue) and genotype-by-sex interaction (orange). Chromosome X is displayed as chromosome 23.

## Results

### Sex-stratified GWAS and genotype-by-sex interaction analysis

We performed sex-stratified GWAS of AD in five cohorts and defined sex by chromosomal composition, with XX individuals classified as female and XY individuals as male. After harmonized sample-level quality control and removal of cross-cohort overlap, the European-ancestry discovery analysis included 449,335 participants: 259,298 females and 190,037 males (**Supplementary Data 1**). The discovery sample comprised 178,948 AD cases and 270,387 controls, with effective sample sizes of 259,298 in females and 190,037 in males. Replication analyses included 230,987 participants from two non-overlapping independent cohorts, comprising 173,644 participants of European ancestry and 57,523 participants of East Asian ancestry. Genotype- and identifier-based screening detected no remaining overlap between the discovery and replication datasets.

The female and male GWAS identified eight and three independent genome-wide significant SNPs, respectively, that were classified as novel under the study criteria (**Supplementary Data 2-3**). The X-chromosome signals comprised rs182269003 in males and rs12689435 and rs5923518 in females; these variants were reported as not previously associated with cognition phenotypes. The sex-stratified results were genetically correlated with the largest sex-combined AD GWAS (female versus both sexes, rg = 0.87 ± 0.03 (rg ± standard error); male versus both sexes, rg = 0.90 ± 0.06) (**Supplementary Fig. S2**, **Supplementary Data 4-5**). Attenuation ratios indicated that most test-statistic inflation was attributable to polygenicity rather than residual population stratification (**Supplementary Fig. S3**). We evaluated 34 female and 22 male independent genome-wide significant SNPs in the independent European HUNT and MVP cohorts. Effect directions were concordant for 85.0% of female signals (95% CI: 70.9–92.9%; H0: concordance = 0.5; *P = 4.18E-06*) and 73.1% of male signals (95% CI: 53.9–86.1%; H0: concordance = 0.5; *P = 0.014*) (**Supplementary Data 6-7**). The 95% confidence intervals were wide, indicating limited precision in the replication analysis.

We next used a genome-wide G×S interaction meta-analysis to test whether SNP effects differed between females and males. The analysis combined the same five cohorts and identified 24 SNPs at genome-wide significance (*P < 5E-08*) and 166 independent SNPs at the suggestive threshold (*P < 1E-06*) (**Supplementary Fig. S3**, **Supplementary Data 8**). There was little evidence of residual population stratification (**Supplementary Fig. S3**). Results for the X-chromosome non-pseudoautosomal region were similar under full and no dosage compensation (**Supplementary Fig. S3**); additional sex-heterogeneity results are reported in **Supplementary Note 1**.

### STAGE-AD integrates sex-aware GWAS and regulatory annotations

Building on DeepGWAS^14^, which models genetic associations with a multilayer neural network, we developed STAGE-AD as a Transformer-based post-GWAS prioritization model. Each variant was represented as an observation combining GWAS summary statistics and population-level genetic metrics with multi-tissue QTLs expression, pathological annotation, functional annotations included chromatin accessibility, enhancer–promoter interaction maps and single-cell transcriptomic profiles from AD brains. The resulting feature space linked 9,034,538 variants to 19,839 candidate genes, 53 tissue or regulatory compartments and 17 cell types. STAGE-AD used these features in a hierarchical multi-token representation to prioritize AD-associated variants and their candidate sex-, tissue- and cell-dependent regulatory contexts (**Supplementary Methods 1**).

To address challenges in defining true binary labels for variant associations, we used a two-step labeling strategy based on GWAS datasets with different statistical power. Association labels were derived from a large AD GWAS, whereas input features were obtained from a smaller independent GWAS with lower power. STAGE-AD did not perform hypothesis testing or generate association P values; which re-ranked SNPs according to their predicted relevance to AD. Among the 131 loci ranked highest by STAGE-AD, 86 mapped to established AD regions and 45 were classified as potentially novel or previously unresolved (**Fig. 1b**, **Table 1**). Despite moderate conventional association evidence, the latter were prioritized through integration of sex-conditioning feature. The model was therefore evaluated as a probabilistic post-GWAS prioritization framework rather than as a replacement for conventional association testing.

STAGE-AD replaced the multilayer perceptron used by DeepGWAS with multi-head self-attention to model nonlinear dependencies among statistical and functional features. We assessed pairwise feature contributions by comparing validation-set changes in area under the receiver operating characteristic curve (AUROC) and area under the precision–recall curve (AUPRC) after permuting individual features and feature pairs (**Supplementary Fig. S4a**). The largest non-additive contributions involved GWAS statistical features, whereas regulatory and QTL annotations contributed mainly marginal information. Removing GWAS statistical features produced the largest performance reduction (AUROC = 0.904 | AUPRC = 0.823). Performance was also lower after excluding position features (AUC = 0.925 | AUPRC = 0.872), molecular QTLs (AUROC = 0.938 | AUPRC = 0.880) or regulatory/open-chromatin-region annotations (AUROC = 0.926 | AUPRC = 0.871). These ablations indicate that GWAS-derived features supplied most predictive information, with functional annotations providing additional discrimination among variants with similar association evidence.

### STAGE-AD prioritization performance

We trained STAGE-AD to model variant-level association statistics, sex-interaction features, LD, fine-mapping probabilities, cross-tissue QTL evidence, epigenomic annotations, regulatory annotations and cell-specific regulatory profiles. The model jointly predicted variant priority, sex-biased locus class, target gene, tissue, cell type and candidate regulatory module. In the test set, the full model achieved an AUROC of 0.947 (95% CI: 0.942-0.952), an AUPRC of 0.895 (95% CI: 0.877-0.912) and a macro-averaged F1 score of 0.847 (95% CI: 0.803-0.891) for variant prioritization and sex-class assignment (**Fig. 2**, **Supplementary Data 11**). The reported Brier score was 0.008 and the expected calibration error was 0.016 (**Supplementary Data 17**).

**Fig. 2.**
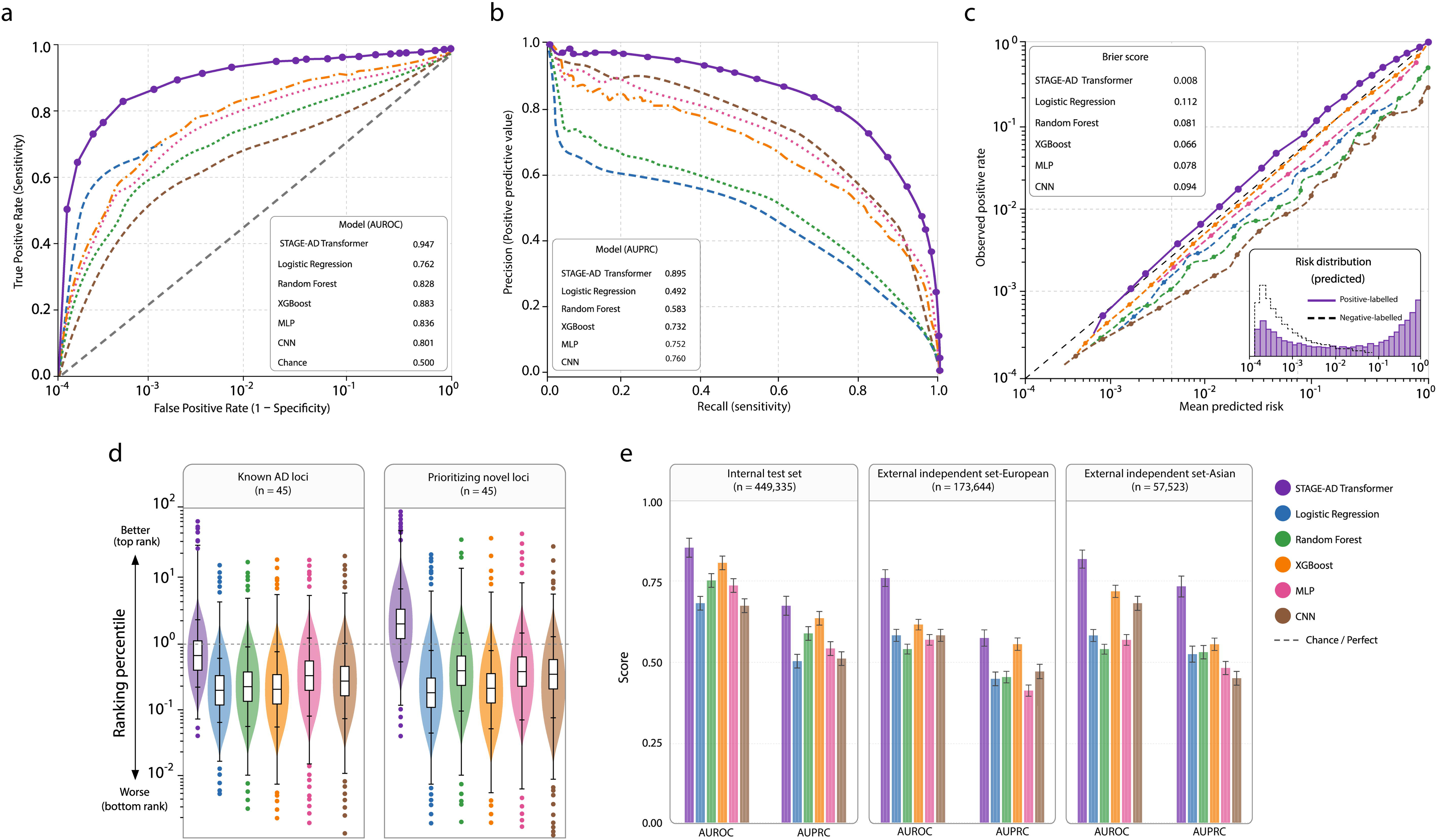
Discrimination, calibration and locus-ranking performance of STAGE-AD. **a,** Receiver operating characteristic (ROC) curves comparing STAGE-AD with logistic regression, random forest, XGBoost, a multilayer perceptron (MLP) and a convolutional neural network (CNN). **b,** Precision–recall curves for the same models, with areas under the precision–recall curve (AUPRC) reported in the inset. **c,** Calibration plots comparing the observed positive rate with the mean predicted probability on logarithmic axes. The black dashed identity line represents perfect calibration. Model-specific Brier scores are reported in the upper-left inset; lower values indicate smaller mean squared prediction errors. The lower-right inset shows predicted-score distributions for the groups labelled ‘Positive-labelled’ and ‘Negative-labelled’ variants. **d,** Distributions of ranking percentiles for previously reported AD loci and candidate novel loci (*n* = 45 per group). Violin plots show the distributions for each model, with embedded box-and-whisker plots. The vertical axis is logarithmic, and the accompanying arrows indicate the ranking direction used in the figure. The horizontal dashed line marks a ranking-percentile value of 1. **e,** Comparison of AUROC and AUPRC across the internal evaluation and external European- and Asian-ancestry datasets. External evaluations used HUNT and MVP for European ancestry and Biobank Japan and the National Biobank of Korea for East Asian ancestry. Throughout, purple, blue, green, orange, pink and brown denote STAGE-AD, logistic regression, random forest, XGBoost, MLP and CNN, respectively.

Relative to the strongest non-Transformer baseline, STAGE-AD increased AUPRC by 16.3% and macro-averaged F1 by 9.7% (**Fig. 2**). The largest reported gains occurred for the underrepresented female-biased, male-biased and G×S-interaction classes; gains were smaller for shared susceptibility variants. Variants in the highest decile of the priority score were enriched 5-fold for genome-wide or suggestive AD association relative to variants in the remaining deciles (*P = 8.63E-22*). The highest decile was also enriched for fine-mapped variants with posterior inclusion probability (PIP) ≥ 0.50 (odds ratio (OR) = 2.84; 95% CI, 1.43–5.62; *P = 0.003*), sex-interaction signals classified as replicated (OR, 2.16; 95% CI, 1.13–4.12; *P = 0.018*) and brain QTL colocalization (OR, 1.92; 95% CI, 1.06–3.47; *P = 0.031*).

### Sex-conditioning and regulatory features contribute to model predictions

We used integrated gradients^15^, attention attribution, feature masking and token-level perturbation to quantify the information domains contributing to STAGE-AD predictions. Variant-level genetic evidence, tissue-specific regulatory annotations and cell-type-specific transcriptomic and epigenomic annotations had the largest aggregate attributions, although their relative contributions differed among locus classes.

Predictions for shared loci were influenced most strongly by expression-associated genetic signals, genetic determinants of protein abundance and chromatin accessibility. Female-biased predictions had larger attributions from female-specific GWAS effects, chromatin accessibility, microglial and astrocytic annotations, blood or immune QTLs and hormone-related regulatory features. Male-biased predictions had larger attributions from male-specific GWAS effects, oligodendrocyte and white-matter annotations. SHAP^16^ analysis ranked APOE (rs429358) as the leading feature among the top 10 features shown in **Fig. 3**.

**Fig. 3.**
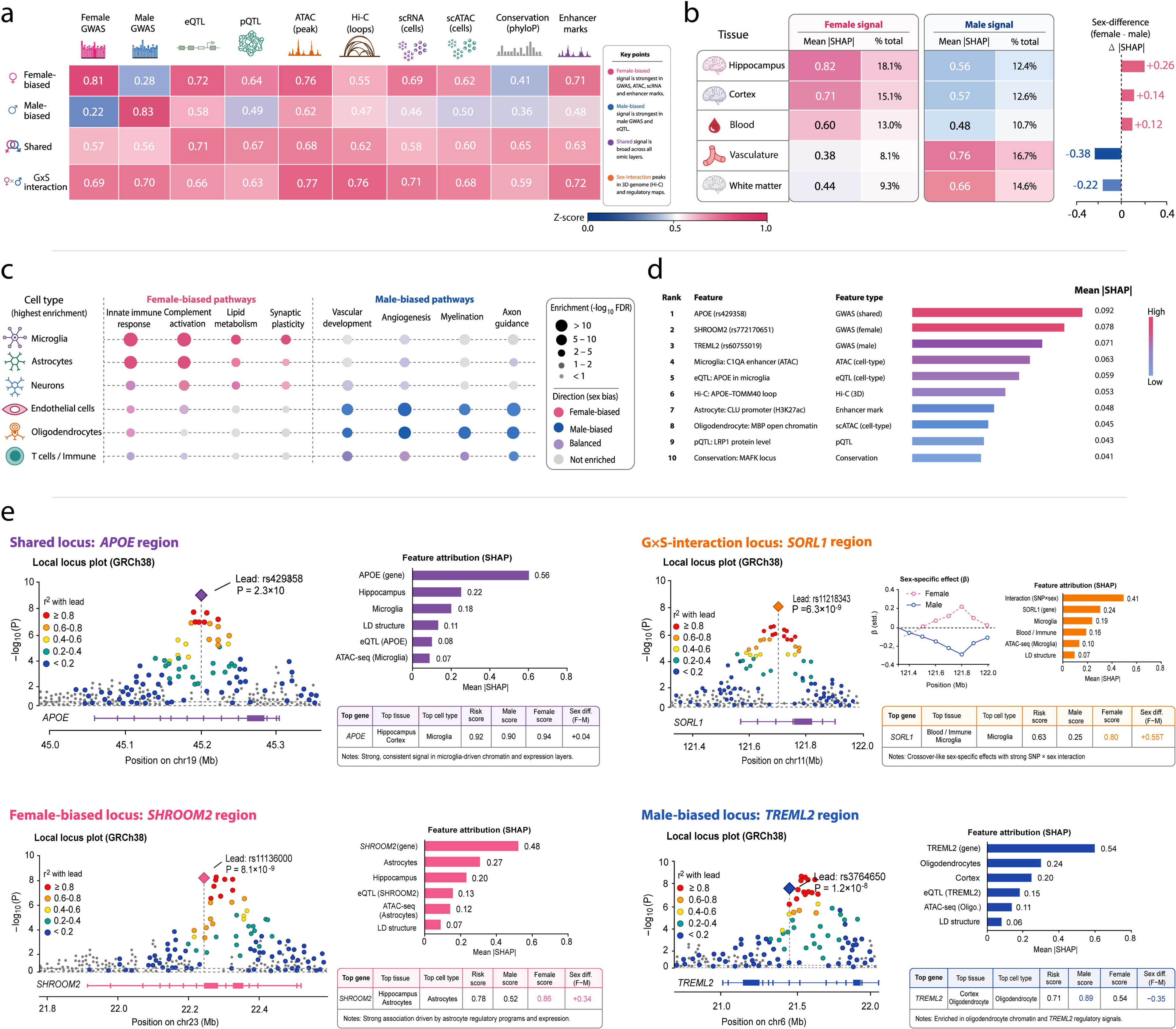
Sex-context-dependent feature attribution and regulatory prioritization by STAGE-AD. **a,** Heat map of genetic and functional feature scores across female-biased, male-biased, shared and genotype-by-sex (G×S) interaction classes. Columns represent female and male genome-wide association study (GWAS) statistics, expression and protein quantitative trait loci (eQTLs and pQTLs), chromatin-accessibility peaks, Hi-C loops, single-cell RNA-sequencing and chromatin-accessibility annotations, sequence conservation and enhancer marks. Cell labels show the displayed *z* scores, with colors ranging from blue for lower scores to pink for higher scores. **b,** Tissue-level SHAP attribution for female and male predictions across the hippocampus, cortex, blood, vasculature and white matter. Tables show mean absolute SHAP values (mean |SHAP|) and percentage contributions. The adjoining bars show female-minus-male differences in mean |SHAP|; positive values indicate greater attribution in female predictions and negative values indicate greater attribution in male predictions. **c,** Cell-type–pathway enrichment profiles across microglia, astrocytes, neurons, endothelial cells, oligodendrocytes and T-cell/immune populations. Pathways encompass innate immune responses, complement activation, lipid metabolism, synaptic plasticity, vascular development, angiogenesis, myelination and axon guidance. Dot size represents enrichment significance as −log₁₀(false-discovery rate (FDR)), grouped according to the displayed size key. Pink, blue, purple and grey indicate female-biased, male-biased, balanced and non-enriched entries, respectively. **d,** Ten displayed features ranked by mean |SHAP|. Feature labels identify the associated variant, gene or regulatory annotation, and the adjacent column indicates the feature type. Bar length and color encode attribution magnitude, with larger values indicating greater contributions to model predictions. **e,** Regional association and feature-attribution summaries for representative shared (upper left), G×S-interaction (upper right), female-biased (lower left) and male-biased (lower right) loci. Regional plots show −log₁₀(*P*) against genomic position on GRCh38, with diamonds marking the lead variants and colored points indicating LD (*r*²) with the corresponding lead variant. Gene tracks are shown beneath each plot. Adjacent bar plots display mean |SHAP| for selected genetic, tissue and cell-type features. Summary tables report the prioritized gene, tissue and cell type, together with model-derived risk-priority scores, male and female scores, and their female-minus-male difference. The *SORL1* inset displays standardized sex-stratified effect estimates across the interval, with pink dashed and blue solid lines representing female and male estimates, respectively.

Removing the sex-conditioning token reduced macro-averaged AUROC for locus classification from 0.947 to 0.767, with the largest reduction for male-biased variants (**Supplementary Data 12**). Macro-averaged AUPRC decreased from 0.895 to 0.778, with the largest reduction for female-biased variants (**Supplementary Data 13**). In this ablation analysis, macro-averaged F1 decreased from 0.847 to 0.751, again with the largest reduction for female-biased variants (**Supplementary Data 14**). Perturbing the sex token changed the predicted class or priority of 16% of candidate loci, indicating that sex-context features contributed information beyond overall GWAS significance.

Leave-one-tissue-out analysis showed the largest reduction in female-biased locus performance after removal of hippocampal information, whereas removal of vascular endothelial annotations produced the largest reduction for male-biased predictions. Masking microglia tokens preferentially reduced female-specific priority scores, whereas masking endothelial or oligodendrocyte tokens preferentially affected male-specific predictions (**Supplementary Fig. S5**).

### Benchmarking and genomic structure-aware sensitivity analyses

We compared STAGE-AD with convolutional neural network (CNN), logistic-regression, XGBoost and multilayer perceptron (MLP) models using consistent input features and AD datasets (**Supplementary Methods 1**). The MLP comparator contained 14 fully connected layers with ReLU activation. Across the reported AUROC, AUPRC and calibration analyses, STAGE-AD ranked above the conventional comparators (**Fig. 2**, **Supplementary Data 11**). Bootstrap resampling was used to evaluate the stability of these comparisons (**Supplementary Fig. S4b**), and the reported calibration advantage was greatest in low false-positive-rate regions. These experiments evaluated predictive discrimination and calibration, not biological validity.

To establish reproducible performance estimates, we benchmarked the proposed model against 13 alternative statistical, machine-learning and deep-learning approaches using identical locus-level training, validation and test partitions. All models were evaluated on the same held-out loci, and all preprocessing and hyperparameter selection procedures were restricted to the training and validation sets. AUPRC was prespecified as the primary performance metric because validated regulatory variants represented a small fraction of the evaluation set. The full sex-conditioned Transformer consistently outperformed association-only and conventional machine-learning approaches. Across the four prediction tasks, the model achieved a macro-averaged AUROC of 0.947 and AUPRC of 0.895, compared with AUPRCs of 0.669 for GWAS P-value ranking, 0.687 for GWAS-only logistic regression, 0.713 for LASSO, 0.724 for random forest and 0.740 for XGBoost. The corresponding AUPRCs were 0.752 for the MLP, 0.760 for the CNN and 0.776 for the standard Transformer (**Supplementary Data 12-13**). Across ranking thresholds, STAGE-AD consistently outperformed the benchmark and ablated models. Precision reached 0.640, 0.526 and 0.441 at the top 100, top 500 and top 1%, respectively, with corresponding recall values of 0.068, 0.230 and 0.397. At the top 1%, precision/recall was 0.457/0.413 for female-biased, 0.422/0.381 for male-biased, 0.467/0.417 for shared and 0.418/0.378 for sex-interaction predictions, demonstrating robust performance across sex-aware prediction tasks (**Supplementary Data 15-16**). The benefit of explicit sex-conditioning was most evident for sex-dependent genetic effects, and the sex conditioning gain cannot be fully explained by the larger sample size of female GWAS. Removing sex-conditioning reduced AUPRC from 0.952 to 0.762 for female-biased variants, from 0.827 to 0.791 for male-biased variants, and from 0.853 to 0.729 for G×S interaction variants (**Supplementary Data 13**). The difference in the interaction task was supported by locus-level paired bootstrap analysis (ΔAUPRC=0.024, 95% CI 0.011–0.037, P<0.001) (**Supplementary Data 18**). Calibration was similarly improved, with an expected calibration error of 0.016 and a calibration slope of 0.99 (**Supplementary Data 17**). Decision-curve analysis further demonstrated the highest prioritization net benefit across prespecified probability thresholds. Together, these analyses indicate that the performance advantage of the full model extends beyond discrimination to candidate ranking, calibration and downstream prioritization utility.

In paired context-model experiments, the sex-conditioned model outperformed sex-blind, permuted-sex and constant-context models for female-biased, male-biased and SNP-by-sex interaction tasks, with smaller gains for shared-risk prediction (**Supplementary Data 19**). The pattern persisted after matching GWAS effective sample size, case–control ratio, minor allele frequency, imputation quality, annotation coverage and missingness, and after excluding the APOE region, chromosome 19 and hormone-related annotations (**Supplementary Data 20-21**). None of 1,000 sex-label permutations matched the full model’s performance (empirical *P < 0.001*) (**Supplementary Data 22**); these analyses did not attribute the gain solely to APOE/chromosome 19 signals. Leave-one-label-out and leave-one-functional-annotation-out analyses indicated that no single label layer or functional annotation class accounted for the full reported performance (**Supplementary Data 23-25**).

To assess genomic leakage, we compared chromosome-stratified LD-block hold-out with SNP-level random partitioning, LD-block, connected-component and strict connected-component hold-out designs (**Supplementary Data 26**). Variants and linked regulatory entities within an LD block were assigned to the same partition, and connected blocks were merged before splitting. AUPRC was 0.895 under chromosome-stratified LD-block hold-out, 0.751 under locus hold-out, 0.811 under chromosome hold-out and 0.681 under APOE-region hold-out evaluation (**Supplementary Data 27**), showing that performance depended on partition design. Under locus hold-out, at least one independently supported variant ranked in the top 1% for 60.5% of held-out loci, and the supported target gene ranked among the top five candidates for 72.8% of loci (**Supplementary Data 28**). Under known-AD-locus hold-out, AUPRC was 0.743 and at least one supported variant ranked in the top 1% for 62 of 85 excluded loci (**Supplementary Data 27,29**). Excluding the APOE region or chromosome 19 reduced AUPRC to 0.681 and 0.774, respectively (**Supplementary Data 27,30**). Notably, APOE remained among the top three candidate genes even when all chromosome 19 variants were excluded from the input data (**Supplementary Data 31**). These structure-aware evaluations support partial transfer to unseen regions while showing that chromosome-stratified LD-block partitioning gave more optimistic performance estimates.

### STAGE-AD nominates candidate AD loci

PLINK clumping at r^2^ greater than 0.10 yielded 118 independent GWAS signals, including all 73 significant signals in the previously reported GWAS summary statistics. The 45 variants in low LD (r^2^ < 0.10) with previously reported AD variants were therefore treated as candidate novel signals under the study definition (**Fig. 1b**, **Table 1, Supplementary Data 32-40**). Their annotations included vascular endothelial development, blood–brain barrier and cell-junction regulation, receptor tyrosine kinase–MAPK/PI3K signaling, calcium and Ras/Rap1 signaling, lipid-droplet metabolism and ABC-mediated transport (**Table 1**). The 2026 consensus GWAS meta-analysis reported 91 Tier 1 loci for AD and related dementias, including 16 new European-ancestry loci^3^. In the comparison presented here, the candidate set was described as 45 STAGE-AD novel variants: 33 autosomal variants were eligible for comparison and 11 X-chromosome variants were excluded because the consensus analysis was restricted to autosomes. None of the 33 autosomal variants matched a consensus lead variant or was within 500 kb of one. The closest pair, rs4988235 and the MGAT5 lead variant rs7557129, was separated by 1.54 Mb; no pairwise LD estimate was available in the 1000 Genomes Phase 3 CEU reference and the pair was not treated as an LD-supported overlap. Thus, these predictions represent non-overlapping candidate regions that require independent genetic and functional evaluation. Differences in phenotype definition, variant representation and analytical strategy may contribute to the lack of overlap. Additional candidate genes are listed in **Supplementary Data 32-35**.

We evaluated the 45 candidate novel variants in independent HUNT^17^ and MVP^18^ summary statistics for two AD phenotypes: (1) self-reported AD (21,635 cases and 57,628 controls) and (2) ICD-based AD diagnosis (31,624 cases and 62,577 controls). Despite differences in phenotype definitions and ascertainment across datasets, 39 variants showed replication in at least one of the two independent European AD cohorts, and 23 loci replicated in both (**Table 1**, **Supplementary Data 32-35**). Six STAGE-AD-prioritized variants (rs10498633, rs144071327, rs183232376, rs4575098, rs772170651 and rs748571391) were also reported in a European AD meta-analysis (GCST90704646-GCST90704648; EADB, UKBB, ADGC, CHARGE, EADI, FinnGen, GERAD, GR@ACE/DEGESCO and PGC-ALZ)^3^, including rs772170651 near SHROOM2, which showed consistent effects in the replication dataset. Because that meta-analysis included UKBB and EADB samples used in the primary analysis, this comparison provides partially independent supporting evidence rather than fully independent replication. Taken together, these results provide additional support for the validity of many of the variants prioritized by STAGE-AD, further reinforcing the method’s ability to uncover robust AD-associated genetic signals that generalize across diverse populations and study designs.

### Statistical-genetic evidence supports STAGE-AD-prioritized loci and genes

Expression- and splicing-based TWAS, conditional and joint multiple-SNP (COJO), fine-mapping and cross-tissue colocalization analyses in HUNT and MVP supported 236 unique genes at 106 SNP-defined loci spanning 86 cytoband regions. At least two approaches supported 105 of 123 STAGE-AD-prioritized SNPs (85.4%), including 39 of 45 candidate-novel SNPs (86.7%) (**Fig. 4a**). Across 125 evaluated SNPs, STAGE-AD rank correlated with the strongest study-defined evidence tier (Spearman’s ρ = 0.724, *P = 1.35E-21*) (**Fig. 4b**). TWAS–QTL directional concordance was recorded for 44 of 57 testable entries (77.2%), representing 54 unique SNPs (**Fig. 4c-e**). Concordance was 11/11 in the highest score decile and 33/46 among lower-scoring entries (*Two-sided Fisher’s exact P = 0.043*). This comparison was descriptive because some SNPs contributed multiple entries. Candidates supported by fewer than two approaches were retained as hypotheses (**Supplementary Data 41**).

**Fig. 4.**
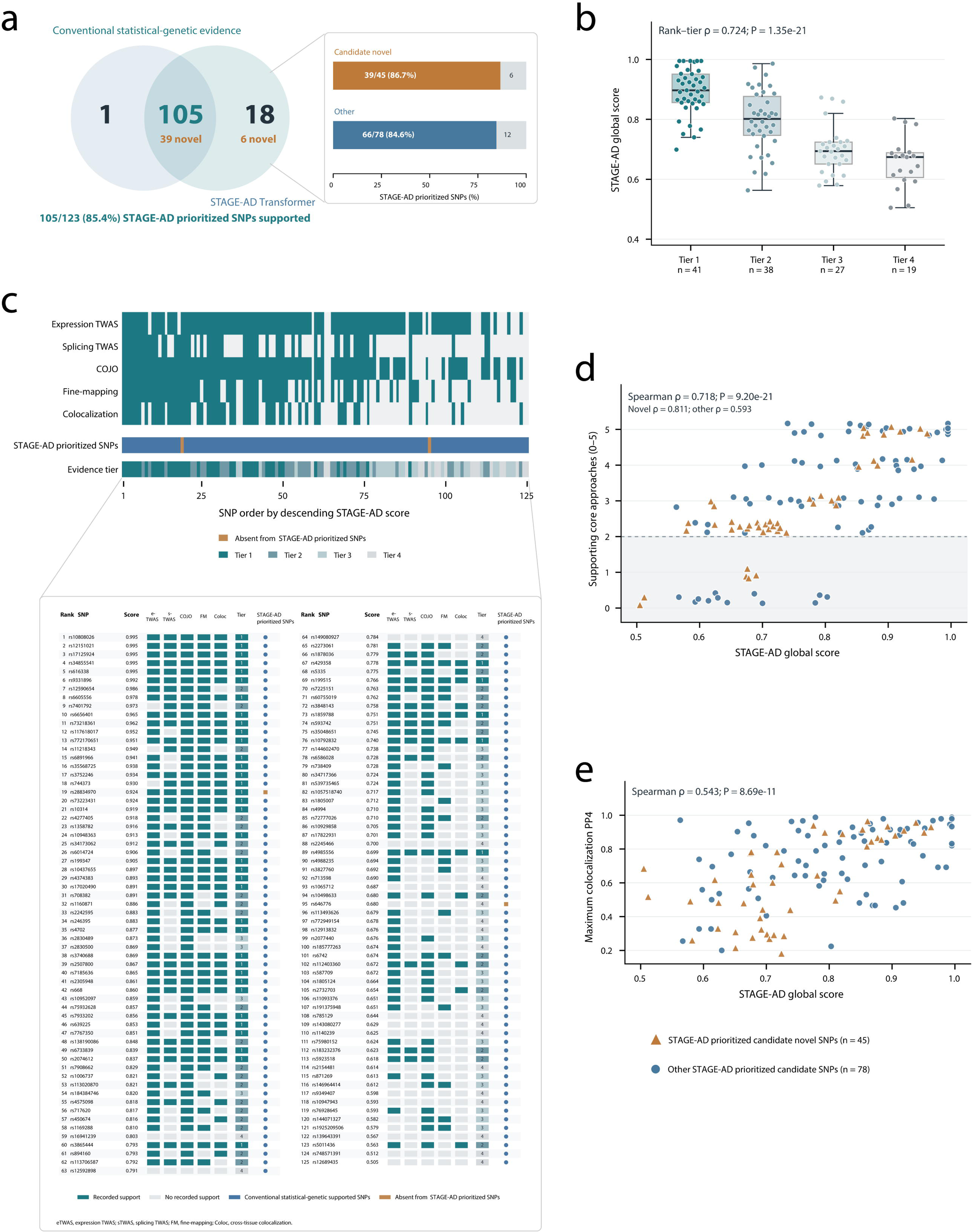
Concordance between STAGE-AD prioritization and statistical-genetic evidence. **a**, Overlap between STAGE-AD-prioritized SNPs and loci supported by at least two statistical-genetic approaches in the independent European HUNT and MVP cohorts. Of 123 prioritized SNPs, 105 (85.4%) had such support, including 39 of 45 candidate-novel SNPs (86.7%) and 66 of 78 other SNPs (84.6%). One supported SNP was not prioritized by STAGE-AD. **b**, STAGE-AD global scores across four integrated evidence tiers for 125 evaluated SNPs (Tier 1, n = 41; Tier 2, n = 38; Tier 3, n = 27; Tier 4, n = 19); Tier 1 denotes the strongest evidence. Points represent SNPs, center lines medians and boxes interquartile ranges. STAGE-AD rank was associated with evidence tier (Spearman’s ρ = 0.724, P = 1.35E-21). **c**, Evidence matrix ordered by decreasing STAGE-AD score. Filled cells indicate support from expression-based or splicing-based transcriptome-wide association studies (TWAS), conditional and joint multiple-SNP analysis (COJO), fine-mapping or cross-tissue colocalization. The tracks below indicate STAGE-AD prioritization and evidence tier; the expanded table lists SNP-level results. **d**, Number of supporting approaches versus STAGE-AD global score for the 123 prioritized SNPs (Spearman’s ρ = 0.718, P = 9.20E-21). The dashed line marks the threshold of two approaches. **e**, Maximum cross-tissue colocalization posterior probability for a shared variant (PP4) versus STAGE-AD global score (Spearman’s ρ = 0.543, P = 8.69E-11). In **d** and **e**, orange triangles denote candidate-novel SNPs (n = 45) and blue circles denote other prioritized SNPs (n = 78).

#### AD susceptibility genes identified for sex-stratified subtypes in TWAS

Sex-stratified expression- and splicing-based multi-tissue TWAS identified 474 unique genes across 248 loci at Bonferroni thresholds of *P < 3.80E-06* and *P < 2.60E-06*, respectively. These results comprised 559 variants and 653 SNP–gene pairs (**Supplementary Data 42**). Under the study’s prior-reporting criteria, 200 genes across 77 loci (42.19% of all unique genes) were classified as not previously reported in AD (**Supplementary Data 42-43**). Six genes across six loci, represented by eight sex-context-specific records, were at least 1.5 Mb from the nearest AD GWAS index variant (**Supplementary Data 42**). Equal-tissue-weight sensitivity analysis largely recovered the genes identified by the brain-upweighted analysis; the few uniquely detected genes had P values near the Bonferroni thresholds. Sex differences in GWAS associations were also evaluated for the 653 SNP–gene models involving 559 variants and 474 genes (**Supplementary Data 43**).

#### Conditioning TWAS and gene-based fine-mapping

After conditioning eQTL and sQTL effects on nearby AD GWAS loci, 41 genes at 25 loci remained at the expression and splicing thresholds (*ep < 3.8E-6* and *sp < 2.6E-6*), excluding APOE. Reported examples included PTK2B (rs4236673), CRHR1 (rs199503) and ARHGAP27 (rs113568679). Gene-level fine-mapping additionally prioritized 67 genes at 31 loci with PIP ≥ 0.9 for at least one eQTL or sQTL in at least one of the 19 tissues (**Supplementary Data 42**). At 11p11.2, a 21-variant credible set was led by rs55743056 at MADD (PIP > 0.92); rs1421155854 overlapped ACP2 and chromatin-interaction data linked rs61915439 to NDUFS3. At 17q21.31, rs530555010 at GRN (PIP = 0.907) was linked to WNT3 (rs199498) in neurons, hippocampus and cortex.

#### Cross-tissue colocalization and functional annotation

Cross-tissue colocalization identified 160 loci at which the AD association and at least one molecular QTL met the predefined posterior-probability threshold for a shared variant. Of these, 77 loci colocalized with brain eQTLs or sQTLs, 49 with blood or immune-cell QTLs and 71 across more than one tissue compartment (**Supplementary Data 44**). Reported female-biased contexts included whole blood, frontal cortex (BA9), anterior cingulate cortex (BA24) and hippocampus; reported male-biased contexts included coronary artery, lung, whole blood and brain neurons. Shared loci were assigned to cortical, microglial and peripheral immune contexts. These distributions are compatible with sex-differentiated regulatory contexts but do not establish tissue mediation (**Supplementary Note 2**).

### Sex-specific genetic architecture of AD

#### Genetic architecture

We estimated sex-specific autosomal SNP-based heritability (h^2^_SNP_), polygenicity (π) and the selection parameter (S) using SBayesS^19^ and the sex-stratified meta-analysis. Liability-scale h^2^_SNP_ assumed lifetime population risks of 0.2 in females and 0.1 in males. Under these assumptions, the posterior probability that female h^2^_SNP_ exceeded male h^2^_SNP_ was 100% (female, 11.42% (95% highest posterior density interval (HPDI): 10.31–12.53%); male, 9.23% (8.28–10.18%)) (**Supplementary Fig. S6a**), consistent with a larger reported h^2^_SNP_ estimate in females under the specified risks. The posterior probability that polygenicity was higher in females was also 100% (female π = 0.044 (0.035–0.056); male π = 0.023 (0.014–0.037)) (**Supplementary Fig. S6b**). Univariate MiXeR^20^ estimated that 17,485 variants (SE = 1680) explained 90% of AD h^2^_SNP_ in females, compared with 17,403 variants (SE = 930) in males (**Supplementary Data 45**). Evidence for a lower selection parameter in males was less certain (82% PP; male S = −0.15 (−0.32–0.13); female S = −0.06 (−0.16–0.09)) (**Supplementary Fig. S6c**), with both intervals including zero.

#### Genetic correlation

Linkage Disequilibrium Score Regression (LDSC) genetic correlations were used to compare sex-stratified AD effects. The genetic correlation of the female GWAS reported by Deming et al.^21^ was higher with the present female GWAS than with the present male GWAS (Z = 4.49, *p-adjust [Benjamini-Hochberg] = 7.3E-06*). The male GWAS reported by Blokland et al.^22^ was more strongly correlated with the present male GWAS than with the female GWAS (Z = 3.23, *p-adjust (Benjamini–Hochberg) = 0.003*). Comparison with Fan et al.^23^ showed a similar but non-significant pattern (**Supplementary Data 4–5**). The female–male genetic correlation was below one (rg = 0.90 ± 0.03; H0: rg = 1; Z = −2.96; *p-adjust [Benjamini–Hochberg] = 0.009*) (**Supplementary Fig. S6d**). Pearson correlations of standardized effect estimate for established AD lead SNPs were additionally evaluated between sexes and across pairwise combinations of the five cohorts (**Supplementary Fig. S6e**).

#### Polygenic overlap

Bivariate MiXeR^20^ reported 17,266 variants (SE = 1850) shared between female and male AD, with an additional 219 variants (SE = 32) unique to females and 137 variants (SE = 34) unique to males (**Supplementary Fig. S6f**). The reported correlation of effect sizes among shared variants was −0.160 (SE = 3.19E-08) (**Supplementary Data 46**). We also accounted for the differential power across our sex-stratified GWAS by repeating MiXeR analyses using equivalent sample sizes in females and males and found the same pattern in European cohorts (HUNT and MVP) (**Supplementary Data 47**). Under gwas-pw^24^ posterior models, 127 genomic regions comprised 120 regions shared between sexes, three female-specific regions and four male-specific regions (**Supplementary Fig. S6g**). Across the 127 regions, 63 candidate variants were mapped to genes by at least two methods (**Supplementary Data 48**), and no additional LD variants were identified in the three female-specific or four male-specific regions (**Supplementary Data 49**). All 120 shared regions showed association peaks in both sexes (**Supplementary Fig. S7**), and the 56 candidate variants in these shared regions had concordant effect directions. These analyses indicate extensive genetic sharing alongside a smaller number of sex-differentiated regions.

### Female-biased loci are enriched for lipid-responsive glial programs

Genes assigned to female-biased and female-enhanced loci were enriched after multiple-testing correction for cholesterol homeostasis (GO:0042632; GO Biological Process), phagocytosis (hsa04145; KEGG), innate immune system (R-HSA-168249; Reactome) and Alzheimer disease (hsa05010; KEGG) (**Supplementary Data 50-51**). The annotated genes involved lipid uptake and efflux, microglial phagocytosis, astrocyte inflammatory reactivity, endolysosomal function and tau-associated neurodegeneration (**Fig. 5a**, **Fig. 3**).

**Fig. 5.**
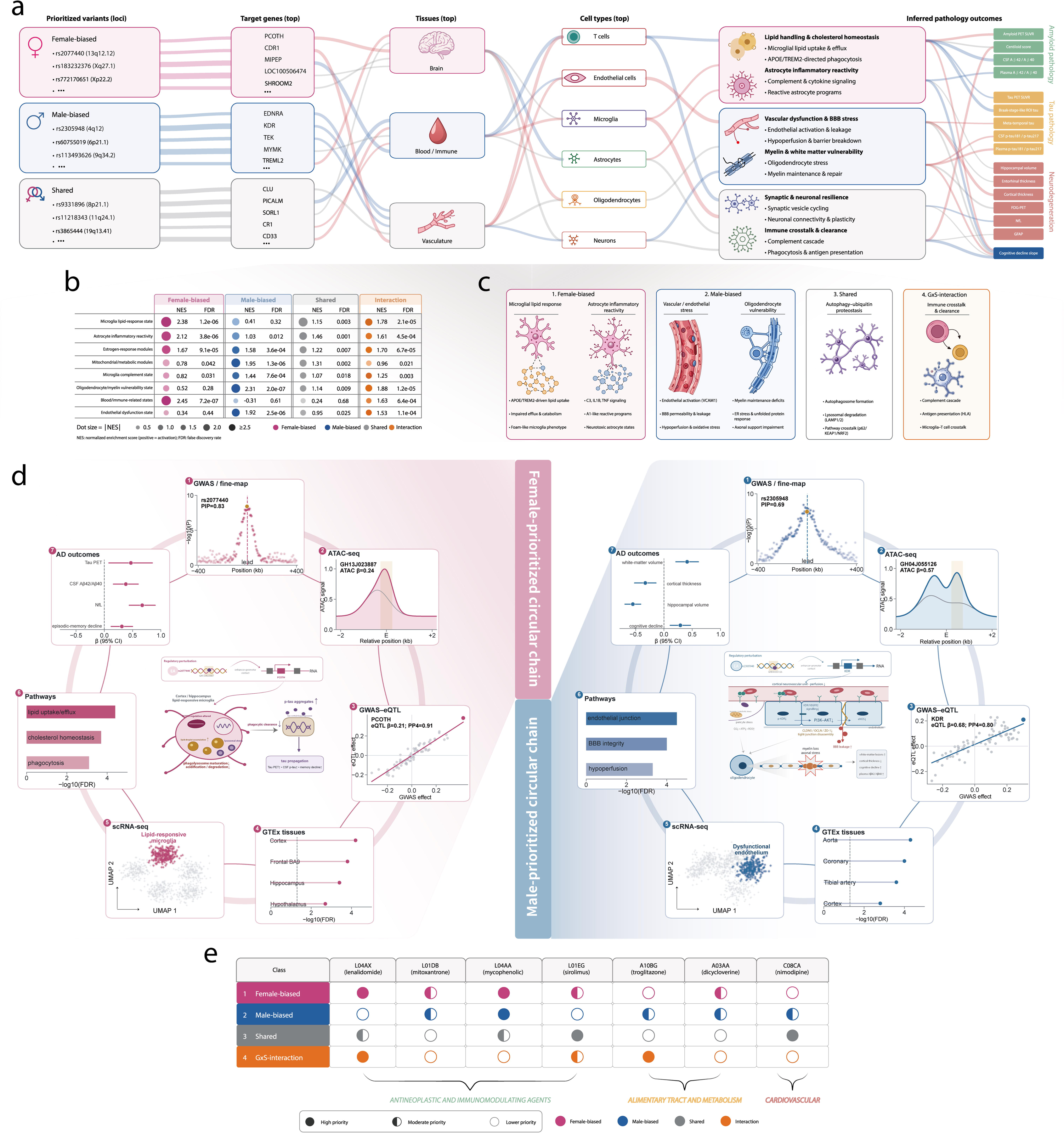
Integrated regulatory and phenotypic characterization of STAGE-AD-prioritized loci. **a,** Alluvial schematic linking selected female-biased, male-biased and shared Alzheimer’s disease (AD) loci to candidate target genes, tissues, cell types, regulatory programs and AD-related phenotypes. Phenotypes are grouped into amyloid pathology, tau pathology and neurodegeneration, with cognitive decline shown separately. **b,** Enrichment of cell states and regulatory modules across female-biased, male-biased, shared and genotype-by-sex (G×S) interaction classes. Numerical entries report normalized enrichment scores (NES) and corresponding false-discovery rate (FDR). Dot size represents the absolute NES. Pink, blue, grey and orange denote female-biased, male-biased, shared and G×S-interaction classes, respectively. **c,** Schematic summary of the regulatory programs prioritized within each sex-context class: microglial lipid response and astrocyte inflammatory reactivity for female-biased loci; vascular or endothelial stress and oligodendrocyte vulnerability for male-biased loci; autophagy–ubiquitin proteostasis for shared loci; and immune crosstalk and clearance for G×S-interaction loci. **d,** Integrated evidence for two representative candidate regulatory chains: rs2077440–*PCOTH* in a female-prioritized microglial context (left) and rs2305948–*KDR* in a male-prioritized endothelial context (right). Numbered subpanels show regional GWAS association and fine-mapping evidence (1), chromatin-accessibility profiles measured by ATAC-seq (2), GWAS–eQTL effect comparisons and colocalization evidence (3), GTEx tissue profiles (4), single-cell RNA-sequencing UMAP embeddings highlighting the nominated cellular states (5), pathway enrichment (6) and associations with AD-related phenotypes (7). PIP denotes the fine-mapping posterior inclusion probability; PP4 denotes the colocalization posterior probability for a shared association signal. Tissue and pathway results are displayed as −log₁₀(FDR). In the outcome plots, points and horizontal lines represent regression coefficients and 95% confidence intervals, respectively; vertical dashed lines indicate no association (β = 0). **e,** Relative compound priorities across the four sex-context classes, derived by comparing expression-TWAS signatures with drug-induced transcriptional profiles from the LINCS Connectivity Map. Filled, half-filled and open circles indicate high, moderate and lower priority, respectively; compound groups are indicated beneath the matrix.

Single-cell analyses assigned female-biased genetic effects primarily to blood or immune states, lipid-response microglia, reactive astrocytes and estrogen-response modules (**Fig. 5b**). Reported enrichment was greater in AD-associated or reactive states than in homeostatic states^25,26^, a pattern consistent with context-dependent regulation.

A highly ranked female-associated module linked rs2077440 at 13q12.12 to PCOTH regulation in cerebral cortex and to lipid-response microglia. Its annotations included microglial lipid uptake and efflux, phagocytosis and endolysosomal catabolism, with support from STAGE-AD variant-to-gene prioritization, tissue and cell-state mapping and literature annotation (**Fig. 5**, **Supplementary Data 53**).

In APOE ε4-stratified analyses, the aggregate female-biased module score showed a nominal interaction for tau-PET standardized uptake value ratio, with a larger reported association among female ε4 carriers than among male carriers or non-carriers (*P-interaction = 0.031*) (**Supplementary Data 52**). The model adjusted for age, ancestry, disease stage and amyloid burden. Estrogen-response and menopause-associated metabolic annotations contributed to five female-biased loci (**Supplementary Data 53**). These annotations identify regulatory regions that may vary across female ageing and endocrine transitions; they do not demonstrate direct hormonal effects.

### Male-biased loci are enriched for vascular and myelination programs

Genes assigned to male-biased loci were enriched for vascular endothelial function, blood–brain barrier integrity, hypoperfusion response, myelin maintenance, axonal support and mitochondrial or metabolic stress. The leading reported enrichments were myelination (GO:0042552; GO Biological Process; false-discovery rate (FDR) = 2.2E-04), cellular response to hypoxia (R-HSA-1234174; Reactome; FDR = 6.8E-04) and fluid shear stress and atherosclerosis (hsa05418; KEGG; FDR = 1.5E-03) (**Supplementary Data 54-55**).

Cell-type analyses assigned male-biased loci preferentially to oligodendrocytes, vascular endothelial cells, complement-associated microglia and astrocytes (**Fig. 5b**). Relative to the female-biased modules, the reported male-biased annotations were concentrated more strongly in vascular and white-matter-associated programs than in lipid-responsive glial states.

At rs10948363 (6p12.3), the prioritized variant colocalized with CD2AP regulation in cerebrovascular endothelial cells and overlapped a regulatory element active in barrier-stressed endothelial cells (**Supplementary Data 56**). The associated evidence chain also included predicted CD2AP regulation, blood–brain barrier integrity, cerebral hypoperfusion, white-matter injury, brain endothelial-cell expression, endothelial regulatory-element overlap, TWAS and experimental blood–brain barrier phenotyping^27^. A second male-associated module linked WNT3, MAPT, NCK2, SEC61G and HS3ST5 to oligodendrocyte, myelin and axonal-support annotations (**Fig. 5c**, **Supplementary Data 54**). Its aggregate score was associated with white-matter hyperintensity volume and processing-speed and executive-function measures, with a reported module-by-sex interaction (*P-interaction = 0.006*).

### Sex-aware genetic modules are associated with AD phenotypes

We tested associations between genetically defined regulatory modules and AD phenotypes in the Alzheimer’s Disease Neuroimaging Initiative (ADNI), Australian Imaging, Biomarkers and Lifestyle study (AIBL), Knight Alzheimer Disease Research Center (Knight ADRC) and Religious Orders Study/Memory and Aging Project (ROS/MAP) (**Supplementary Data 57**). The female microglial–lipid and astrocyte-reactivity module was associated with plasma p-tau217, CSF p-tau181, tau positron-emission-tomography burden and episodic-memory decline (**Fig. 5 Fig. 1a**, **Supplementary Data 58-59**), with the largest reported estimates among women carrying APOE ε4. The male vascular–myelination module was associated with white-matter lesion burden, cortical thickness, hippocampal or cortical atrophy and processing-speed and executive-function decline (**Supplementary Data 58**, **Supplementary Data 62**). Module-by-sex interaction analyses indicated differences in longitudinal cognitive-decline slopes after multiple-testing correction (**Supplementary Data 59**, **Supplementary Data 62**). Shared autophagy–ubiquitin proteostasis, synaptic and neuronal resilience, and immune crosstalk and clearance modules were associated with amyloid and tau phenotypes in both sexes (**Fig. 5a**, **Supplementary Data 58**, **Supplementary Data 62**), although the downstream tissue and cell-state annotations differed. Associations were reported to remain similar after adjustment for APOE-related pathology (**Supplementary Data 62**).

In neuropathological datasets, 3 of 4 modules were associated with Braak stage, CERAD score, quantitative amyloid and tau burden. Directions were consistent across 3 independent cohorts and remained similar after exclusion of individuals with mixed or non-AD neuropathology (**Supplementary Data 60**). Mediation analysis estimated that tau-PET burden accounted for 31% of the association between the female microglial–lipid and astrocyte-reactivity module score and episodic-memory decline (**Supplementary Data 61**).

### Integrated evidence prioritizes candidate regulatory chains and potential drug candidates

We integrated GWAS z score, G×S-interaction z score, fine-mapping PIP, QTL colocalization, epigenomic annotation, tissue and cell-type enrichment, protein-level expression, model attribution, and biomarker and neuropathology associations into locus-specific evidence chains (**Supplementary Data 63**). These candidate regulatory chains were evaluated in the independent individual-level Neuroscience Project–Alzheimer’s Disease (NPAD) cohort.

The integration identified 11 high-confidence genetic–tissue–cell type–pathology chains (**Fig. 5d**, **Supplementary Data 64-68, Supplementary Fig. S9**). A female-prioritized chain linked rs2077440 at 13q12.12 to PCOTH regulation in hippocampal and cortical tissue, lipid-responsive microglia, tau-PET burden and episodic-memory decline. A male-prioritized chain linked rs2305948 to KDR regulation in cerebrovascular endothelium, barrier-stressed endothelial-cell dysfunction, blood–brain barrier breakdown, white-matter lesion burden and cortical atrophy. Mendelian randomization analyses were treated as supplementary statistical evidence for directional consistency rather than as experimental validation (**Supplementary Data 69-75**).

Drug-repurposing analyses produced distinct rankings for female-biased, male-biased, shared and genotype-by-sex interaction modules (**Fig. 5e**, **Supplementary Fig. S10**). For female-biased modules, lenalidomide and mycophenolic acid were top-ranked, whereas mitoxantrone, sirolimus and dicycloverine received moderate priority. For male-biased modules, mycophenolic acid was top-ranked, with mitoxantrone, troglitazone, dicycloverine and nimodipine receiving moderate priority. Shared modules ranked sirolimus and nimodipine highest and assigned moderate priority to lenalidomide and mycophenolic acid. The genotype-by-sex interaction module ranked lenalidomide and troglitazone highest, with sirolimus receiving moderate priority. Immunomodulatory compounds were represented most often among female-biased and genotype-by-sex interaction rankings, whereas the male-biased and shared rankings also included metabolic and cardiovascular drug classes. These results are hypothesis-generating predictions from genetic-module associations and do not establish therapeutic efficacy, clinical safety or treatment direction.

## Discussion

In this study, we integrated sex-stratified and genotype-by-sex association analyses with cross-tissue molecular and cellular evidence to characterize sex-dependent components of AD susceptibility within a predominantly shared genetic architecture. Analyses of 449,335 participants of European ancestry identified eight female and three male genome-wide-significant signals provisionally classified as novel, together with 24 variants showing genome-wide-significant genotype-by-sex interactions. We developed STAGE-AD to connect these association patterns to candidate variants, target genes and regulatory contexts through sex-conditioned integration of molecular QTLs, epigenomic features and single-cell annotations. The model improved prioritization over association-only and alternative machine-learning approaches, with ablation and matched-context comparisons supporting the contribution of sex conditioning, although performance decreased under more stringent genomic hold-outs. Among 45 prioritized candidate variants, 39 received support in at least one independent European cohort, and complementary statistical-genetic analyses supported 236 genes at 106 loci. Genetic correlation and polygenic-overlap analyses indicated extensive sharing between sexes alongside a smaller component of effect heterogeneity. Tissue and cell-state analyses implicated female-biased microglial lipid responses and astrocyte reactivity, male-biased endothelial and oligodendrocyte programs, and shared proteostatic, synaptic and immune processes. Associations with biomarkers, neuroimaging, cognition and neuropathology linked these modules to disease-related phenotypes, including tau burden and episodic-memory decline for female-prioritized modules and white-matter injury and executive-function decline for male-prioritized modules. Integrated evidence prioritized 11 candidate regulatory chains, which were evaluated using multi-omics and neuropathological data from the independent NPAD cohort. Transcriptomic signature analyses further nominated drug-repurposing candidates across sex contexts. Collectively, these findings suggest that deep learning framework may be crucial for fully harnessing existing datasets and achieving a more comprehensive understanding of AD genetic susceptibility, showcasing the value of identifying potential causal genes, functional pathways, and drug targets for AD prevention.

The sex-stratified results are more consistent with predominantly shared AD susceptibility than with two distinct genetic architectures. The female–male genetic correlation was 0.90 ± 0.03, 120 of 127 gwas-pw regions were assigned to a shared model, and MiXeR estimated similar numbers of variants explaining 90% of SNP heritability in females and males. This pattern accords with a previous sex-stratified analysis reporting a genetic correlation of 0.96 and similar SNP-heritability estimates across sexes^28^. The present data nevertheless support a limited heterogeneous component, including formal genotype-by-sex signals and modest deviation of the cross-sex genetic correlation from one. Such effects are plausible given reported age-dependent APOE associations and sex-dependent genetic associations with AD endophenotypes^29^. The modest number of X-linked findings is also consistent with recent large AD X-chromosome analyses, which identified relatively few genome-wide signals despite substantially larger samples and emphasized the analytical and power constraints specific to chromosome X^8,9^. These observations should, however, be separated from differences in statistical detection. The female discovery sample was larger than the male sample (259,298 versus 190,037 participants), and the case distributions also differed. Down-sampling, matching and permutation analyses reduce, but cannot eliminate, power-related explanations for the larger number of female signals and the higher performance of the female-biased prediction task. Accordingly, significance in one sex and non-significance in the other was not treated as evidence of a sex difference unless supported by the genotype-by-sex interaction test, consistent with current recommendations for sex-aware genetic analyses^30^.

Phenotype heterogeneity is another important source of both generalizability and uncertainty. The contributing cohorts used clinical diagnoses, electronic health-record codes, self-report or family-history proxies, and different follow-up structures. Proxy AD can be genetically correlated with clinically diagnosed disease, as shown by the reported genetic correlation of 0.81 in a large AD-by-proxy meta-analysis^31^, but recent work has demonstrated that parental-history designs can be affected by survival and participation biases^32^. Such differences can dilute association estimates, alter class labels used for model training and change calibration across cohorts. In the present study, 39 of 45 prioritized candidate variants received support in at least one of HUNT or MVP and 23 received support in both, despite differences in ascertainment. This constitutes cross-cohort corroboration rather than uniform replication of an identical phenotype. The additional European meta-analysis comparison was only partially independent because it included datasets used in discovery. Clinical-only, proxy-excluded and cohort-specific sensitivity analyses therefore remain important for determining which signals are stable across definitions and which are phenotype dependent.

The comparison with simpler models clarifies the incremental value of the Transformer. With identical partitions and input features, STAGE-AD achieved an AUPRC of 0.895, compared with 0.669 for GWAS P-value ranking, 0.687 for GWAS-only logistic regression, 0.740 for XGBoost, 0.752 for the multilayer perceptron, 0.760 for the convolutional neural network and 0.776 for a standard Transformer. Sex conditioning was particularly informative for the interaction task, for which the paired locus-level improvement in AUPRC was 0.024 (95% confidence interval, 0.011–0.037; *P < 0.001*). These comparisons suggest that the gain arose from the combination of structured molecular annotations, contextual interactions and explicit sex conditioning rather than model depth alone. This interpretation is consistent with post-GWAS neural-network approaches that integrate association statistics with functional annotations^14^ and with a recent Transformer analysis of migraine, which also concluded that curated features accounted for much of the performance gain^33^. The structure-aware evaluations are equally informative: the reduction in AUPRC under locus, chromosome and APOE-region hold-outs indicates partial, not complete, transfer to unseen genomic contexts. Similarity-aware splitting is necessary in biological machine learning because random or weakly structured partitions can yield optimistic estimates when related observations occur across partitions^34^. STAGE-AD should therefore be viewed as a post-GWAS ranking method that concentrates follow-up effort, not as a substitute for association testing or as evidence that a high-ranked variant is disease causing.

The biological annotations suggest different emphases within a largely shared architecture. Female-weighted predictions were enriched for lipid handling, phagocytosis, innate immunity, lipid-responsive microglia and reactive astrocytes. These patterns are compatible with the immune and lipid pathways identified by large AD GWAS^2^ and with human multi-omic studies reporting sex-dependent microglial immunometabolism^35^. Experimental and single-cell work has also described sex- and APOE-dependent neutrophil–microglia states, providing a plausible context in which immune effects may vary between females and males^36^. Male-weighted predictions emphasized vascular endothelium, blood–brain barrier regulation, myelination, hypoxia and white-matter injury. Cerebrovascular burden and its cognitive consequences show sex-dependent patterns in AD cohorts, although the direction varies by vascular exposure, brain region and outcom^37^. The present enrichment and ablation results therefore refine candidate cellular contexts; they do not demonstrate that glial processes are exclusive to females or vascular and oligodendrocyte processes exclusive to males.

Convergence across fine-mapping, TWAS, colocalization, QTL direction, neuropathology and module–phenotype associations increase the priority of selected chains, but convergence remains distinct from experimental validation. TWAS estimates genetically predicted expression associations, colocalization evaluates compatibility with a shared statistical signal, mediation models rely on identification assumptions, and model attributions quantify which inputs influence predictions. None alone establishes the sequence of molecular events from variant to pathology. Attention weights in particular are not intrinsically faithful explanations of model behavior^38^, and integrated gradients remain model-dependent attributions rather than biological perturbations^15^. The 11 candidate chains should therefore be tested by editing the prioritized allele or regulatory element in isogenic human induced-pluripotent-stem-cell models. Cell types should match the nominated context, including microglia, astrocytes, brain endothelial cells and oligodendrocytes, and perturbations should be evaluated in relevant co-cultures or blood–brain barrier organoids. Prespecified readouts should include target-gene expression, chromatin accessibility or contact, and the predicted cellular phenotype, followed where appropriate by amyloid, tau, phagocytosis, lipid-handling, barrier-integrity or myelination assays. Single-cell CRISPR interference and prime editing have already provided a workable route from AD association signals to experimentally supported microglial regulatory effects^39^. Replication in independent human brain single-cell multi-omic data and, for the strongest chains, orthogonal organoid or in vivo models would further reduce cell-state and model-system dependence.

The drug-repurposing results require an even higher evidential threshold. Lenalidomide has reached a phase II proof-of-mechanism trial design in amnestic mild cognitive impairment due to AD, but the published report describes the protocol rather than efficacy^40^. Sirolimus is supported by preclinical rationale, whereas a small open-label phase I study of rapamycin in ten participants detected no drug in cerebrospinal fluid, found no significant cognitive change and reported increases in several AD-related and inflammatory biomarkers^41^. A Cochrane review of nimodipine found some short-term benefit for cognition and global impression but no benefit for activities of daily living and insufficient evidence to justify long-term use^42^. Mitoxantrone reduced Aβ seeding and pathology in cellular and APP-transgenic mouse experiments^43^, but separate mouse data showed neuronal injury and tau hyperphosphorylation^44^. Troglitazone reduced tau phosphorylation in a cell model^45^ but was withdrawn from the US market because of hepatotoxicity^46^. Dicycloverine (dicyclomine) is an anticholinergic, and a systematic review identified cognitive impairment with dicyclomine exposure^47^, arguing against direct prioritization in an AD population. We did not identify direct human AD efficacy evidence for mycophenolic acid. Thus, the drug rankings are best used to identify perturbable pathways or chemically testable signatures. Clinical candidacy would additionally require evidence for direction of effect, brain exposure, dose, target engagement and safety, followed by randomized evaluation.

Several features strengthen the study. The analysis used a large discovery sample, formal genotype-by-sex tests, explicit X-chromosome modeling, non-overlapping external cohorts and multiple definitions of replication. Model comparisons used common inputs and partitions, evaluated AUPRC and calibration in addition to AUROC, and included ablations, label permutations and increasingly stringent genomic hold-outs. Biological interpretation did not depend on a single annotation layer: candidate loci were evaluated using independent statistical-genetic methods and downstream molecular and pathological data. Negative or attenuated findings, including the loss of model performance under strict hold-outs and the imprecision of some replication estimates, are informative because they define where the framework is most and least reliable.

The study also has limitations. Discovery was restricted to European-ancestry participants, and many LD, QTL and regulatory resources used for training are European enriched. Evaluation in East Asian cohorts is useful but does not demonstrate portability to African, admixed or other underrepresented populations, nor does it remove ancestry dependence in allele frequency, LD, molecular-QTL effects or calibration. Multi-ancestry AD studies have shown that diverse LD structures can refine loci and reveal heterogeneous effects^48^, and European-derived AD polygenic scores show variable transfer across populations^49^. Future versions should be retrained and calibrated using ancestry-matched LD and molecular-QTL panels, include local-ancestry analyses in admixed samples, and report performance within each ancestry rather than only in pooled data. Residual phenotype misclassification, unequal female and male sample sizes, cohort-specific ascertainment, and the assumed lifetime risks used to transform SNP heritability to the liability scale also affect interpretation. Because model labels were derived from higher-powered GWAS and most predictive information came from GWAS features, performance may partly reflect recovery of familiar association structure. Finally, postmortem molecular data are vulnerable to disease-stage effects and cannot determine temporal order; statistical mediation, drug-signature reversal and model attribution do not resolve this limitation. The absence of direct perturbational validation remains the principal barrier to mechanistic claims.

In conclusion, our findings indicate that the genetic architecture of AD is predominantly shared between females and males but includes a smaller component of sex-differentiated genetic and regulatory effects. By integrating sex-stratified and genotype-by-sex association signals with cross-tissue molecular QTLs, epigenomic annotations and disease-relevant cellular states, STAGE-AD prioritizes candidate variants, genes and regulatory contexts beyond those resolved by conventional association analyses alone. The resulting female-weighted glial and lipid-response programs and male-weighted vascular and oligodendrocyte programs provide testable hypotheses for how shared AD genetic risk may be differentially manifested across biological contexts. These results establish a framework for translating sex-aware genetic association signals into experimentally tractable regulatory hypotheses and refine our understanding of the genetic architecture underlying sex differences in AD. Looking ahead, incorporating sex-specific genetic insights into clinical practice may enable more refined approaches to risk assessment and therapeutic stratification in AD. These results may inform the development of novel therapeutics that target sex-dependent biological mechanisms, ultimately improving precision medicine strategies.

## Methods

### Study populations, phenotype definitions and sex assignment

Discovery analyses used individual-level genotype and phenotype data from five international cohorts and included 449,335 participants of genetically inferred European ancestry. The cohorts were the Alzheimer’s Disease Genetics Consortium (ADGC)^50^ (female: cases = 12,516, controls = 8,128; male: cases = 9,102, controls = 10,025), the Alzheimer’s Disease Sequencing Project (ADSP)^51^ (female: cases = 10,239, controls = 15,209; male: cases = 11,961, controls = 18,013), the European Alzheimer & Dementia Biobank (EADB)^2^ consortium (female: cases = 11,788, controls = 12,562; male: cases = 8,924, controls = 9,883), UK Biobank (UKBB)^52^ (female: cases = 55,194, controls = 72,211; male: cases = 29,308, controls = 50,686) and All of Us^53^ (female: cases = 19,788, controls = 41,663; male: cases = 10,128, controls = 32,007). The external independent European cohorts included HUNT^17^ (female: cases = 14,207, controls = 24,994; male: cases = 8,917, controls = 13,172) and MVP^18^ (female: cases = 19,094, controls = 50,151; male: cases = 11,221, controls = 31,888), which were performed by STAGE-AD Transformer validation and conventional statistical-genetic analyses. The external independent Asian cohorts included BBJ^54^ (female: cases = 5,126, controls = 6,027; male: cases = 3,260, controls = 5,884) and NBK (female: cases = 8,763, controls = 10,165; male: cases = 6,263, controls = 12,035), which were only utilized by STAGE-AD Transformer cross-ancestry validation. All contributing studies complied with the relevant ethical regulations and obtained informed consent from participants. Cohort-specific information is provided in **Supplementary Methods 1**; **Supplementary Data 76** summarizes implementation of the recommendations for sex-aware analyses described by Khramtsova et al.^30^.

AD cases were defined within each cohort using available combinations of electronic health records, hospital diagnosis codes, death-registry records, medication records, cognitive assessments, clinically adjudicated diagnoses and neuropathological confirmation. Controls had no recorded AD, dementia, mild cognitive impairment or related neurodegenerative diagnosis during the available observation period (**Supplementary Data 1**).

Where parental-history proxy phenotypes were available, they were analyzed separately and were not combined with clinically diagnosed AD in the primary analysis unless otherwise specified. Sensitivity analyses excluded proxy cases and restricted the phenotype to clinically diagnosed AD. Prespecified exclusions comprised missing sex information, discordance between genetically inferred and recorded sex, sex-chromosome aneuploidy, poor genotyping quality, close relatedness beyond the allowed threshold and ancestry-outlier status.

In this study, sex refers to genetically inferred or registry-recorded biological sex. Genetically inferred sex was used when genotype-based information was available.

### Genotyping, ancestry inference, quality control and imputation

Cohort-specific genotyping platforms, quality-control procedures and imputation methods are described in **Supplementary Methods 1**. Genetic ancestry was inferred by principal-component analysis of linkage-disequilibrium-pruned common variants after exclusion of high-LD and long-range-LD regions. Individual-level quality control in each European cohort excluded participants outside the European-ancestry analysis group, whereas Asian cohorts only remained the Asian-ancestry participant.

### Sex-stratified GWAS and genotype-by-sex interaction analyses

Association analyses were conducted within each cohort using fastGWA^55,56^ in GCTA v1.94.1. Sex-stratified GWAS of the binary outcome used a mixed linear model (--fastGWA-mlm-binary), a sparse genetic relationship matrix, the first 10 ancestry principal components and any cohort-specific covariates. Autosomes and the X chromosome were analyzed. Genome-wide G×S analyses used a mixed linear model (--fastGWA-mlm), the same form of genetic relationship matrix, sex as the environment variable, the first 10 ancestry principal components and any cohort-specific covariates. The G×S analysis included the autosomes and the X-chromosome non-pseudoautosomal region under no dosage compensation (--dc 0) and full dosage compensation (--dc 1). Sex-stratified and G×S results retained SNPs with minor allele frequency larger than 0.01 and r^2^ imputation score larger than 0.6; cohort-specific details are in **Supplementary Methods 1**. Sex-stratified summary statistics were used to estimate sex-specific SNP effects and for downstream SNP-based heritability, genetic-correlation and polygenic-overlap analyses. The G×S analysis formally tested differences in SNP effect estimates between sexes.

For each variant, retained fields comprised the overall, female-specific, male-specific and G×S-interaction effect estimates; standard errors; z scores; P values; effect-allele frequency; and sample size. Variants were assigned to shared, female-biased, male-biased, G×S-interaction or uncertain classes using the prespecified criteria based on sex-specific effect estimates, directional concordance, sex-stratified P values and G×S-interaction evidence.

Genome-wide significance was defined as *P < 5E-08* and suggestive significance as *P < 1E-06*. Both thresholds were evaluated for G×S interactions. Interpretation required consistency between the interaction result and the sex-stratified effect estimates and did not rely on the interaction P value alone.

### APOE-region and APOE-stratified analyses

The APOE region was analyzed separately because of its effect size, linkage-disequilibrium structure and reported sex-differentiated associations with AD risk and tau pathology^7^. The locus was defined as chromosome 19:44–46 Mb on GRCh38. APOE ε2, ε3 and ε4 haplotypes were inferred from rs429358 and rs7412 when the variants were directly genotyped or confidently imputed.

Primary analyses were run with and without adjustment for APOE ε4 dosage. Sensitivity analyses excluded the APOE region, stratified participants by APOE ε4 carrier status and tested the G×S-by-APOE interaction. COJO^57^ analyses were combined with the TWAS by conditioning on the APOE region to evaluate the dependence of reported sex-related signals on APOE.

### Meta-analysis and locus definition

Cohort-specific GWAS summary statistics were combined by inverse-variance-weighted fixed-effect meta-analysis. Random-effects meta-analysis was also performed for loci with evidence of between-cohort heterogeneity. Genomic inflation was evaluated using genomic-control lambda and the LD score regression intercept^58^.

The five-cohort sex-stratified and G×S results were inverse-variance-weighted meta-analyzed. The source methods reported 109,525 cases and 149,773 controls in females and 69,423 cases and 120,614 controls in males. Because cohort-level G×S analyses used linear mixed models, beta estimates were converted to the logistic scale by first converting beta values to OR with the R function described by Lloyd-Jones et al.^59^ and then taking log(OR). The standard error of log(OR) was calculated as log(OR) divided by the Z score. Standard-error-weighted meta-analyses without genomic control were conducted in METAL^60^ (release 05/05/2020). Meta-analysis results retained SNPs present in three or more cohorts (at least 50% of the cohorts). Lead independent SNPs were identified by PLINK v1.90b6.8^61^ clumping with --clump-r^2^ = 0.1 and --clump-kb = 1000.

Independent loci were defined by linkage-disequilibrium clumping of genome-wide significant and suggestive variants. Within each locus, the variant with the smallest P value was selected as the lead. Locus boundaries used either ±1 Mb around the lead variant or LD-based intervals; complex loci, including APOE, were manually adjusted. Overlapping loci were merged when their credible sets or LD structure were considered to represent the same association signal.

### Functional-genomic resources and harmonization

GWAS meta-analysis results were integrated with molecular, regulatory, pathological annotations. Molecular-QTL resources included multi-tissue eQTLs, sQTLs, pQTLs and brain- and immune-specific QTLs from Genotype-Tissue Expression (GTEx)^62^ Project, eQTLGen^63^, PsychENCODE^64^ and AMP-AD^65^. Epigenomic annotations comprised candidate cis-regulatory elements, chromatin-accessibility peaks, histone-modification peaks, ChromHMM states, transcription-factor binding sites and enhancer–gene links from ENCODE^66^, Roadmap Epigenomics^67^, AAD model resources^68^, promoter-capture Hi-C datasets^69^ and brain-specific regulatory maps^70^. AD-related single-cell and single-nucleus resources^71^ supplied cell-type-specific expression, disease-associated expression, sex-biased expression and chromatin-accessibility features (**Supplementary Methods 2**).

All genomic coordinates were harmonized to GRCh38. Variants used allele-specific identifiers in the form chromosome:position:reference:alternate. Ensembl gene identifiers were stripped of version suffixes and mapped to HGNC symbols for presentation. Tissue, brain-region and cell-type names were mapped to a controlled vocabulary. QTL effect alleles were aligned to GWAS effect alleles. Resources on other genome builds were converted by liftOver, and ambiguously mapped variants or elements were excluded.

### Single-cell and single-nucleus feature extraction

Processed single-cell and single-nucleus RNA-sequencing datasets were used preferentially (**Supplementary Methods 2**). Within each dataset, cells or nuclei were grouped by donor, brain region, disease status and cell type, and pseudo-bulk profiles were generated at the donor-by-cell-type level. Gene-level features comprised mean expression, percentage of expressing cells, cell-type-specificity z score, tau-specificity score, AD-associated differential expression, sex-biased expression and AD-by-sex interaction estimates.

Differential-expression models included age, sex, post-mortem interval, brain region, batch, sequencing depth and RNA quality as covariates. Sex-biased expression was estimated within cell type; AD-by-sex interaction models were fitted where sample size permitted. Single-cell chromatin-accessibility data were used to derive cell-type-specific accessible peaks, gene-activity scores and overlaps between variants and regulatory regions. Peaks were assigned to genes by promoter overlap, enhancer–gene maps, chromatin-contact evidence or gene-activity models.

Brain cell types included excitatory neurons, inhibitory neurons, astrocytes, microglia, oligodendrocytes, oligodendrocyte precursor cells, endothelial cells, pericytes and vascular leptomeningeal cells. Monocytes, macrophages, T cells, B cells and natural killer cells were included as complementary immune and peripheral-cell resources relevant to AD.

### Sex-aware AD regulatory graph and candidate instances

A regulatory graph represented relationships within AD-associated loci. Nodes denoted variants, credible sets, regulatory elements, genes, tissues, cell types, sex contexts and AD-pathology states. Edges encoded credible-set membership, variant–gene associations, variant overlap with regulatory elements, enhancer–gene links, chromatin accessibility, cell-type enrichment, sex-biased expression, sex-biased QTL effects and pathology-associated molecular changes.

Candidate variant–gene pairs were formed when a variant had molecular-QTL evidence, overlapped a regulatory element linked to the gene, occurred in a chromatin-contact region involving the gene promoter, mapped to the gene body or promoter, or was assigned by proximity within the locus window. Candidate genes required at least one functional, regulatory or proximity link to a credible-set variant. Each pair was expanded across the relevant tissues, cell types, pathology states and sex contexts to create variant–gene–tissue–cell-type–pathology–sex-context instances.

### Weak-supervision labels, prediction tasks and data partitioning

Because experimentally validated AD variants and genes are incomplete, training used weak supervision. Positive labels were assigned to candidate mechanisms supported by multiple study-defined evidence layers: concordant effect direction, reported AD risk genes, experimentally supported enhancer–gene links, and previous reported pathology evidence. Low-confidence and negative examples were candidate genes in the same locus without QTL support, regulatory links, pathway evidence. Negative labels were down-weighted, and positive-unlabeled learning was used in sensitivity analysis to evaluate false-negative bias.

Sex-context labels were derived from overall, female-biased, male-biased and G×S-interaction GWAS results. Female-enriched labels required stronger genetic evidence in females, concordant effect direction and supporting interaction or heterogeneity evidence; male-enriched labels were defined analogously. Shared labels required concordant effects in females and males without strong interaction evidence. Ambiguous instances were labeled uncertain and were either excluded from supervised sex-class training or assigned lower weights.

The multi-task objective combined gene-prioritization, variant-prioritization, tissue/cell-type classification, pathology-prioritization and sex-context classification losses. Binary cross-entropy, categorical cross-entropy and pairwise-ranking losses were assigned according to the prediction task. Chromosome-stratified LD-block, rather than individual variant–gene pairs, were allocated to training (70%), validation (15%) and test (15%) sets to limit information leakage. Hyperparameters were selected on validation performance, and the final model was evaluated in held-out loci and external datasets.

### Model inputs and token construction

Each model instance represented a candidate regulatory mechanism within an AD-associated locus. Heterogeneous tokens represented the locus, variant, gene, regulatory element, enriched tissue, cell type, pathology state and sex context; numerical and categorical features were assigned to the corresponding tokens.

Variant features comprised overall (shared), female-biased, male-biased and G×S-interaction GWAS beta, SE, z scores; P values; effect-allele frequency (EAF); minor-allele frequency (MAF); CADD score^72^; distance to the transcription start site; APOE-region status; LD score; distance to the nearest gene; distance to TSS; coding consequence; RegulomeDB score^73^; chromatin-state conservation score; and overlap with the GWAS Catalog^74^ (previously reported studies). Gene features comprised QTL statistics; sex-biased expression; gene type and referred nearest loci; eQTL, sQTL and pQTL target-gene assignments; and chromatin-accessibility- and ABC-linked genes^75^. Gene-token attributes included distance to the variant; constraint score; expression in brain, microglia, astrocytes, neurons, oligodendrocytes and endothelial cells; AD differential expression; and sex differential expression. Regulatory-element features comprised chromatin-accessibility state, ATAC signal, histone-modification signal, enhancer class and enhancer–gene score. Tissue and cell-type features encoded biological context. Tissue tokens included GTEx v8 (49 tissue) and BrainSpan^76^ (fetal, childhood, adolescent and adult brain), with tissue-name embedding; eQTL, sQTL and pQTL z scores; tissue-specific expression. Cell-type tokens included microglia, astrocytes, excitatory neurons, inhibitory neurons, oligodendrocytes, OPCs, endothelial cells, pericytes, vascular leptomeningeal cells, choroid-plexus epithelial cells and T cells / myeloid immune cells, with marker enrichment, expression specificity and module-eigengene correlation with pathology states. Sex-context tokens represented overall, female, male and G×S-interaction contexts and included overall, female, male and G×S-interaction variants and z scores; beta_female minus beta_male; the female-to-male OR ratio; APOE ε4 × sex effect; X-chromosome annotation; XCI-escape status; sex-biased gene expression; sex-hormone-pathway annotation; and a menopause-related trait. Final inputs comprised instance-level tables, token files, edge files, token-feature matrices, label files and locus-level train–test split files.

### Sex-aware cross-tissue regulatory Transformer

The model was designed to learn dependencies among variants, genes, regulatory elements, tissues, cell types, pathology states and sex contexts. Token embeddings combined entity-type embeddings, numerical-feature projections, categorical embeddings and context embeddings. Sex embeddings were supplied as explicit tokens and as conditioning variables in the attention layers.

The architecture contained four Transformer encoder layers^77^ with multi-head self-attention, feed-forward layers, residual connections, layer normalization and dropout. Graph-aware attention-bias terms derived from edge types and weights in the regulatory graph were added to encode the specified biological relationships. Full details of feature construction, model architecture, optimization, sensitivity analyses and reproducibility are provided in the **Supplementary Methods 2** and **Supplementary Fig. S1**.

The four primary outputs were a candidate-gene probability, variant-priority score, tissue/cell-type relevance score and sex-context regulatory class. The classes were female-biased, male-biased, shared, G×S-interaction and uncertain. Auxiliary outputs described eQTL-mediated, sQTL-mediated, pQTL-mediated, enhancer-mediated, chromatin-contact-mediated and mixed candidate regulatory mechanisms.

### Model evaluation, comparisons and interpretability

Evaluation metrics were the AUROC, AUPRC, top-k recall of reported AD genes, F1 score, calibration error and locus-level ranking performance. Comparators were model with GWAS P value ranking only, GWAS beta, SE and P value, LR, graph-aware attention bias, LASSO, RF, XGBoost, MLP, CNN, standard Transformer, sex-blind Transformer, full sex-conditioned Transformer (STAGE-AD), No-P Transformer, No-functional annotation Transformer.

Ablations removed sex-condition tokens, variant features, tissue/cell-type features, epigenomic features or graph-aware attention bias to estimate the change in predictive performance attributable to each information domain. Attention weights, integrated gradients, feature attribution and in silico token perturbation were used to describe model sensitivity. For each prioritized locus, the highest-contributing paths were extracted as candidate, rather than experimentally established, regulatory mechanisms.

To reduce the probability that variants and linked regulatory instances from the same LD block entered different partitions, evaluation included random SNP-level partitioning, LD-block hold-out, chromosome-stratified LD-block hold-out, connected-component hold-out based on shared target genes, strict connected-component hold-out based on shared target genes and regulatory elements. Incremental information from the sex context was assessed with paired models using otherwise identical feature inputs: sex-condition (variant-level features + gene-level features + regulatory-element features + tissue/cell-type tokens + sex-context tokens), sex-blind (variant-level features + gene-level features + regulatory-element features + tissue and cell-type tokens), permuted-sex (variant-level features + gene-level features + regulatory-element features + tissue and cell-type tokens + randomly permuted sex-context tokens) and constant-context. Sensitivity analyses down-sampled the female GWAS to the male effective sample size; matched the case/control ratio, MAF and INFO, annotation count and missingness; removed the APOE region, all of chr19 or hormone/menopausal/androgen annotations; and permuted the sex label 1000 times.

### External cohorts and transcriptome-, splice- and protein-level analyses

To explain the new sex specific loci and potential mechanisms proposed by the STAGE-AD framework, we conducted external validation using two independent European-ancestry populations to assess the reliability of the Transformer framework in a biological context. We evaluated the genetic associations of prioritized AD-related genes in two independent population-based cohorts (**Supplementary Methods 1**).

#### Joint-tissue expression and splicing-based TWAS analyses

TWAS analyses were performed using prediction models from GTEx Project^62^ previously built using genotyping and RNA-sequencing data across 49 tissues derived from individuals of European ancestry. In this study, we used models from 19 tissues relevant to AD etiology including brain-specific tissues (Amygdala, Anterior cingulate cortex, Caudate, Cerebellar Hemisphere, Cerebellum, Cortex, Frontal Cortex, Hippocampus, Hypothalamus, Nucleus accumbens, Putamen, Spinal cord, and Substantia nigra), fat tissues (subcutaneous adipose and visceral adipose), immune cell-related tissues (whole blood, Epstein-Barr virus-transformed lymphocytes, and spleen), and the liver.

We applied two joint-tissue TWAS procedures: 1) expression based and 2) splicing based. For the expression analysis, S-PrediXcan^78^ was run separately for each gene and each of the 19 tissues using eQTL models. Gene-level P values were combined across all 19 tissues with the aggregated Cauchy association test (ACAT)^79^, assigning equal weights to non-brain tissues and a combined weight to brain tissues equivalent to all other tissues. For the splicing analysis, S-PrediXcan and sQTL models were first applied to each sQTL within each intron-excision cluster. ACAT then combined sQTL signals within clusters and across the 19 tissues, again weighting brain tissues as equivalent to all other tissues combined. Equal weights across all tissues were examined in sensitivity analysis. Each procedure was applied to overall, female-specific, male-specific and G×S-interaction GWAS summary statistics.

#### Conditioning our TWAS analyses to adjust for nearby GWAS variants

To assess whether expression- and splicing-based TWAS associations were independent of nearby reported GWAS signals, both analyses were repeated after conditioning on genome-wide significant index SNPs (*P < 5E-08*). Effects of SNPs in the eQTL and sQTL prediction models were conditioned on significant GWAS index variants within ± 2 mega-base-pair (Mb) of each gene’s transcription start and stop sites. COJO was used to estimate the conditioned eQTL and sQTL effects, which were then supplied to the expression- and splicing-based TWAS procedures described above.

#### Proteome-wide association study

FUSION^5^ was applied with plasma pQTL resources from HUNT and MVP. Transformer-prioritized genes were compared with pQTL colocalization, protein-level association and AD-brain proteomic-change evidence.

#### Colocalization and molecular QTL validation

To account for correlation among gene–trait associations induced by LD, expression- and splicing-TWAS results for each sex context were fine-mapped with Fine-mapping of Causal Gene Sets (FOCUS)^80^. Analyses supplied FOCUS with subtype-specific summary statistics and an LD matrix computed from GTEx genotype data with bigsnpr^81^, separately for eQTLs, sQTLs and each of the 19 tissues. Within each LD block, 90% credible sets were constructed separately for genes and intron-excision events, and marginal PIPs were calculated for each gene within each of the 19 tissues. Genes with PIP ≥ 0.9 for eQTLs or any sQTLs were designated candidate genes. Sex-context colocalization compared female GWAS–QTL with male GWAS–QTL results; a mechanism was designated female- or male-biased when the colocalization evidence was stronger in that sex and aligned with the corresponding GWAS effect estimate.

### Partitioned heritability and regulatory-annotation enrichment

#### Genetic architecture of AD in females and males

Autosomal SNP-based heritability (h^2^_SNP_), polygenicity (π) and the selection parameter (S), defined here as the relationship between effect size and minor-allele frequency, were estimated from the sex-stratified meta-analysis summary statistics using SBayesS in GCTB v2.5.2^19^. Analyses used the linkage-disequilibrium shrunk sparse matrix of 2.8 million variants and four chains of length 25,000, with burn-in of 5000 and thinning of 10. H^2^_SNP_ was converted to the liability scale by the method of Lee et al.^82^, assuming population prevalence of 0.2 in females and 0.1 in males. For h^2^_SNP_, π and S, the posterior probability that the female value exceeded the male value was the proportion of Markov chain Monte Carlo (MCMC) samples satisfying that inequality. Sensitivity analyses estimated h^2^_SNP_ with LDSC, accounted for differential power between the sex-stratified GWAS, and examined male under-diagnosis and between-cohort heterogeneity. LDSC v1.0.1^83^ with European LD scores from 1000 Genomes estimated bivariate autosomal SNP-based genetic correlations (rg). Correlations compared the sex-stratified summary statistics with the largest European-only sex-combined AD GWAS^3^, previous sex-stratified AD GWAS and the present female and male meta-analyses. To examine between-cohort heterogeneity, rg was also estimated for pairwise combinations of the two sexes across the five cohorts used in the meta-analyses. Sex differences in effect estimates for established sex-combined AD SNPs were also evaluated.

#### Polygenic overlap of AD across sexes

Univariate and bivariate MiXeR v1.3^20^ quantified polygenicity and polygenic overlap between male and female AD. The Gaussian-mixture model represents common-variant effects as a mixture of non-null and null components; polygenicity was reported as the number of model-assigned variants explaining 90% of h^2^_SNP_. Sex-shared and sex-differentiated regions were evaluated with gwas-pw v0.21^24^ using the recommended European BED file of approximately independent LD blocks. Cross-trait correlation was set to zero because the female and male GWAS contained no overlapping individuals. Regions with posterior probability of association (PPA) larger than 0.5 under models one, two and three were assigned to female-only, male-only or shared association models, respectively. In each assigned region, the SNP with the largest PPA above 0.5 was selected as a candidate variant. SNP2GENE in FUMA v1.5.2^84^, ANNOVAR^85^ and Ensembl v110 were used for annotation. Positional mapping used a 10 kb window. EQTL mapping used all tissues from TIGER^86^, InsPIRE^87^, EyeGEx^88^, eQTL catalogue^89^, PsychENCODE^64^, scRNA eQTLs, DICE^90^, eQTLGen^63^, Blood eQTLs, MuTHER^91^, xQTLServer^92^, CommonMind Consortium^93^, BRAINEAC^94^ and GTEx v8. The 3D chromatin-interaction mapping used built-in data comprising Hi-C from 21 tissue/cell types in GSE8711286^95^, Hi-C loops from Giusti-Rodríguez et al.^96^, PsychENCODE Hi-C^64^ and FANTOM5 enhancer–promoter correlations^97^. PsychENCODE, FANTOM5 and Brain Open Chromatin Atlas^70^ annotations were applied to all three mapping procedures. Genes assigned to a SNP by more than one procedure were retained.

### Functional annotation and cell-type enrichment

MAGMA v1.08^98^ in FUMA was used for gene-based, gene-set and gene-property analyses of the sex-stratified and G×S results. Inputs were the summary statistics and PLINK-clumped lead independent SNPs. Settings were Ensembl v110; maximum lead-SNP P value and P-value threshold of 5E-08 for sex-stratified results or 1E-06 for G×S results; first and second r^2^ thresholds of 0.1; the European 1000 Genomes Phase 3 reference panel^99^; and a 10 kb gene window. Gene-set tests used gene-level statistics from all MAGMA-tested genes and 10,678 sets (4761 curated sets and 5917 Gene Ontology terms) from MSigDB v6.2^100^; significance used Bonferroni-corrected P values. Gene-property tests assessed whether genes with greater tissue-specific expression had stronger AD association, using RNA-sequencing data from 30 general and 53 GTEx tissue types and brain samples from 11 general developmental stages in BrainSpan. Genome-wide significant SNPs from the sex-stratified analysis were assigned to genes by positional, eQTL and 3D chromatin-interaction mapping using the procedures described under ‘***Polygenic overlap of AD across sexes.***’

Cell-type enrichment used gene-specificity metrics derived from single-cell data. Enrichment of GWAS signals and Transformer-prioritized genes was tested across the included brain, immune, vascular and metabolic cell types. Female-enriched, male-enriched and shared prioritized gene sets were analyzed separately.

### Associations with AD pathology and biomarkers

Prioritized genes were integrated with AD-related phenotypes from six cohorts: ADNI, AIBL, Knight ADRC, ROS/MAP, ACT and Mayo Clinic Brain Bank (**Supplementary Data 57**). Available outcomes comprised clinical AD diagnosis; cognitive decline; amyloid and tau PET; CSF and plasma biomarkers; and neuropathological traits including Braak stage, neuritic-plaque burden, amyloid burden, tau burden and CERAD score. These datasets were used to test associations between genetically prioritized genes and the measured AD-related phenotypes.

For participant *i*, the raw genetic score for regulatory module *k*, constructed using the sex-stratified discovery GWAS for sex *s*, was calculated as

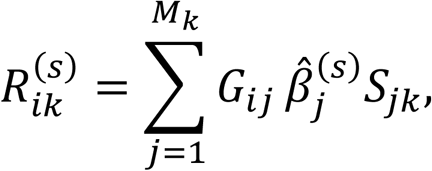

where *M_k_* is the number of LD-independent variants assigned to module *k*, *G_ij_* is the effect-allele dosage for variant *j* in participant *i*, 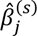 is the corresponding female- or male-stratified GWAS effect estimate, and *S_jk_* is the dimensionless STAGE-AD variant-to-module prioritization score. The Transformer score incorporated evidence linking each variant to its prioritized target gene, tissue and cell state; genes, tissues and cell types were therefore not treated as independent genotype-bearing units in the participant-level score. Effect alleles were harmonized across the discovery GWAS and validation datasets before score calculation. Only LD-clumped variants passing the prespecified quality-control criteria were included.

Raw scores were calculated separately for female- and male-prioritized modules and standardized within each validation cohort:

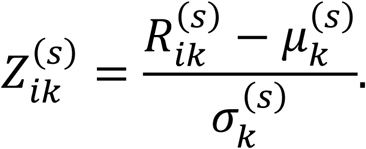

Association coefficients therefore represent the expected change in the outcome per 1-SD increase in the corresponding module score. For analyses formally comparing effects between women and men, module scores were standardized using the pooled cohort mean and standard deviation, and sex dependence was evaluated using a module-score-by-sex interaction term.

Continuous pathology outcomes, including Braak stage^101^, CERAD score^102^ and Thal phase, were analyzed by multivariable linear regression with the stats R package; binary outcomes by logistic regression; and time-to-event outcomes by Cox proportional-hazards models in the survival R package. Models were fitted separately in females and males, and sex differences were tested directly with a molecular-feature-by-sex interaction term. P values were adjusted by the Benjamini–Hochberg FDR procedure. Covariates comprised age, sex, ancestry, APOE genotype, post-mortem interval, brain region and cohort-specific confounders.

### Candidate regulatory cascades and external multi-omics evaluation

Female-enriched and male-enriched candidate cascades integrated Transformer priority, standardized GWAS z scores, QTL colocalization, epigenomic annotation, single-cell expression, protein-level evidence and pathology association. Each cascade linked a variant, regulatory element, candidate target gene, tissue, cell type and AD-related pathological process. Study-defined high-confidence cascades required support from at least five evidence layers; cascades supported by one evidence type were retained as hypotheses.

Candidate cascades were evaluated with multi-omics data from the external NPAD cohort (**Supplementary Methods 3**). Sex-prioritized genetic module scores used audited, LD-clumped independent variants and sex-specific GWAS effect estimates as weights. Participant module scores were the weighted sums of effect-allele dosages and were standardized within cohort. Associations with AD-related pathological outcomes were estimated in each cohort by multivariable regression adjusted for age, sex, ancestry principal components and phenotype-specific covariates. Sex-dependent associations were tested with module-score×sex interaction terms rather than by comparing significance between sex-stratified models. Cohort estimates were combined by inverse-variance-weighted meta-analysis; heterogeneity was assessed with Cochran’s Q statistic and I^2^, and FDR control was applied. Two-sample Mendelian randomization^103^ used prespecified instruments to evaluate directional statistical evidence. Instrument strength was assessed by F statistics; sensitivity analyses used weighted median^104^, MR-Egger regression^105^, horizontal-pleiotropy tests, Steiger directionality^106^, Cochran’s Q and leave-one-out analyses. Statistical mediation models estimated indirect effects with nonparametric bootstrap confidence intervals.

### Drug-repurposing analysis

Following a genetics- and transcriptomics-informed signature-mapping procedure^107^, sex-aware AD expression signatures were constructed from expression-TWAS gene-level Z scores for female-biased, male-biased, genotype-by-sex-interaction and shared components. Drug-induced profiles were obtained from the LINCS Connectivity Map Level 5 replicate-consensus data^108^ after CMap quality control. Disease and drug signatures were compared with an ensemble connectivity score combining the weighted connectivity score, XSum, XCor, XSpe and XCos. Scores were standardized and averaged across the prespecified gene-selection thresholds and score types; negative scores denoted predicted reversal of the AD-associated expression signature. Empirical P values used 100 within-tissue gene-label permutations and a +1 correction. Compounds were ranked by connectivity-score magnitude, consistency across tissues or sex-context signatures and convergence with genetic evidence.

### Sensitivity analyses

Sensitivity analyses included evaluation in external European- and Asian-ancestry datasets; exclusion of the APOE region; adjustment for APOE ε4 dosage; restriction to clinically diagnosed AD; exclusion of proxy cases; and re-training of the Transformer after feature ablation. Additional analyses assessed sensitivity to sex-specific sample-size imbalance, case–control imbalance, allele-frequency differences, phenotype definition, imputation quality, annotation density and missingness. Calibration and ranking stability were evaluated by bootstrap resampling and in external datasets.

### Statistical analysis

Statistical tests were two-sided unless specified otherwise. Genome-wide significance was *P < 5E-08*. The Benjamini–Hochberg FDR was used for multiple-testing control in transcriptome- and splice-TWAS, pathway and cell-type-enrichment analyses. Regression assumptions were evaluated with residual diagnostics and sensitivity analyses. Effect estimates were accompanied by standard errors or 95% confidence intervals. Random seeds were fixed for model training, data partitioning and negative sampling. Intermediate files, model-input tables, hyperparameters, trained checkpoints and analysis scripts were archived for reproducibility.

### Ethical approval and data governance

The study complied with relevant regulations for research involving human participants and the principles of the Declaration of Helsinki. Analyses used de-identified data. Each cohort obtained participant informed consent and approval from the relevant institutional review board or ethics committee (**Supplementary Methods 1,3**). Use of controlled-access genetic, transcriptomic, neuropathological, imaging and biomarker data complied with the applicable data-use agreements.

## Data Availability

The raw genotype and phenotype data used to run the association analyses in all separate cohorts (ADGC, ADSP, EADB, UKBB, All Of Us, HUNT, MVP, BBJ, NBK) are protected and are not available due to data privacy laws. Summary-level results are provided in the Supplementary Data. Publicly shareable sex-stratified and genotype-by-sex GWAS summary statistics, STAGE-AD prioritization results and data dictionaries are available through Zenodo (https://doi.org/10.5281/zenodo.22651873).

Individual-level genotype and phenotype data from the contributing cohorts, including genetic, multi-omic, clinical and neuropathological data from NPAD, are not publicly available owing to data-privacy restrictions. De-identified NPAD data and the associated data dictionary may be requested from the corresponding author. Requests should specify the research objectives, proposed analyses and required datasets, accompanied by relevant ethics documentation. The project director will assess compatibility with participant consent and applicable data-protection requirements. Access requires the relevant approvals and a data-use agreement specifying permitted uses, data-security requirements and restrictions on re-identification and onward disclosure.

## Code Availability

All code used in this study is publicly available at our GitHub repository (https://github.com/zhiangcheng/STAGE-AD-Transformer). To support flexibility and reproducibility, we provide configurable training pipelines, pretrained model checkpoints, and documentation to facilitate comparative analysis and transparent reporting.

## Acknowledgements

We thank the study participants, brain donors and their families, particularly those in the NPAD cohort, and the teams involved in clinical characterization, tissue collection, neuropathological assessment and molecular profiling. We acknowledge the investigators and staff of the Alzheimer’s Disease Genetics Consortium, Alzheimer’s Disease Sequencing Project, European Alzheimer & Dementia Biobank, UK Biobank, All of Us Research Program, HUNT, Million Veteran Program, BioBank Japan and National Biobank of Korea for generating and providing access to the genetic data used in this study. We also thank the investigators and staff of ADNI, AIBL, Knight ADRC, ROS/MAP, ACT and the Mayo Clinic Brain Bank for the biomarker, cognitive and neuropathological resources, and the consortia and research groups that generated and shared the molecular-QTL, epigenomic, chromatin-interaction and single-cell datasets supporting these analyses. This research received no specific grant from any funding agency in the public, commercial or not-for-profit sectors.

## Author contributions

Z.C., S.J. and A.G. conceived and designed the study. Z.C. and S.J. developed STAGE-AD, implemented the computational pipelines and performed the sex-stratified GWAS and genotype-by-sex meta-analyses. S.J. and J.P. curated the functional-genomic datasets and conducted statistical-genetic validation and integrative multi-omics analyses. J.P. and A.G. coordinated the NPAD cohort, including sample collection, clinical characterization, neuropathological assessment and molecular profiling. Z.C. and J.P. prepared the figures and tables. Z.C. and S.J. wrote the initial manuscript, and A.G. supervised the study. All authors contributed to the interpretation of results, critically revised the manuscript and approved the final version. Z.C. and S.J. contributed equally to this work.

## Table Legends

**Table 1** STAGE-AD-prioritized Alzheimer’s disease variants and candidate regulatory contexts.

