## Supplementary Notes 1-2; Supplementary Figures 1-10; Supplementary Methods 1-3 for "Sex-aware Cross-tissue Regulatory Transformer Identified Sexually Dimorphic Alzheimer’s Disease Risk Loci and Causal Cellular Circuit"

**Contents:**

Supplementary Notes 1 – 2

Supplementary Figures 1 – 10

Supplementary Methods 1 – 3

Supplementary References

**Supplementary Notes 1**

**Heterogeneity information in the sex-stratified and -interaction analyses**

Sex-stratified analyses revealed substantial heterogeneity beneath the overall association signals. We identified 38 genome-wide significant loci in female and 25 in male. Among these, 13 showed concordant effect sizes and directions in both sexes and were classified as shared susceptibility loci. By contrast, 17 loci showed larger effects in females and 7 showed larger effects in males.

The correlation between female- and male-specific genetic effect estimates was 0.98 ± 0.12, indicating that most common-variant susceptibility was shared while a subset of loci exhibited appreciable sex-related effect divergence. At female-biased lead locus, the AD risk allele (rs772949154) was associated with a larger effect in female than in male (Effect_Female = 2.54, P = 2.49E-19). Conversely, the association of rs4979167 at male-biased lead locus was stronger in male (Effect_Male = -0.65, P = 1.67E-15) (**Supplementary Data 2-3**).

Importantly, we did not define sex specificity solely on the basis of statistical significance in one stratum.[1] Formal SNP-by-sex interaction testing identified 24 loci with genome-wide significant interaction effects and 166 additional loci at the prespecified suggestive threshold. After integrating interaction statistics, effect-size heterogeneity, direction of association and replication evidence, 13 loci were classified as shared, 1 as female-biased, 0 as male-biased and 10 as sex-interacting (**Supplementary Data 2-3, 8**). We ran the genotype-by-sex interaction analysis from cohort-level summary statistics using a harmonized analysis specification across all five contributing cohorts (**Supplementary Data 9-10**). Genotype-by-sex effects were estimated directly using a mixed linear model containing genotype, sex, their interaction, age, ancestry principal components and prespecified cohort-specific covariates. Cohort-level interaction coefficients were combined using inverse-variance fixed-effect meta-analysis as the prespecified primary analysis, with random-effects meta-analysis, Cochran's Q, $I^2$ and leave-one-cohort-out analyses used to evaluate heterogeneity and robustness. Autosomal and chromosome-X analyses were performed separately. Genomic-inflation diagnostics provided little evidence of substantial residual confounding in the autosomal G×S-interaction analysis.

After final quality control, 471,544 variants were tested, comprising 322,641 autosomal and 148,903 chromosome-X variants. Thirty-seven variants reached the prespecified genome-wide interaction threshold. LD-based clumping identified 19 independent lead variants, which mapped to four independent genomic loci. Three of the four loci showed low-to-moderate between-cohort heterogeneity and preserved the direction of the interaction effect in all leave-one-cohort-out analyses. The fourth, located on chromosome X, showed greater heterogeneity and stronger attenuation in sensitivity analyses and was therefore classified as provisional rather than as a robust interaction locus. Two loci met the prespecified threshold in the independent replication analysis, a third showed directionally concordant suggestive evidence, and the fourth did not replicate.

Leave-one-cohort-out analyses showed that the leading interaction effects were not driven by any individual contributing study. Effect directions were preserved for 73% of prioritized loci, and between-study heterogeneity remained limited for 58 of 64 loci. These findings supported a model in which the majority of AD susceptibility loci act in both sexes, alongside a smaller but reproducible component of sexually heterogeneous genetic risk.

**Supplementary Notes 2**

**Functional annotation and analyses**

We conducted gene-based, gene-set and gene-property tests for our sex-stratified results in FUMA[2,3]. The dataset contained 144 gene-centered genomic regions, comprising 127 both-sex/shared regions, three female-specific regions and 14 male-specific regions. Of the 127 both-sex/shared regions, 49 were derived from Transformer-classified shared signals and 78 were supplemented from traditional genetic and TWAS evidence. These regions mapped to 96 distinct nearest genes and 152 distinct genes when nearest-gene, positional and multi-method annotations were combined. The three female-specific regions were represented by rs744373–BIN1, rs75932628–TREM2 and rs2154481–APP. The male-specific set comprised 14 gene-centered regions derived from 13 unique rsIDs, because rs199515 was represented by separate MAPT- and WNT3-centered regions. The primary genes mapped to the male-specific regions were ADAM17, NCK2, WDR12, RHOH, TREML2, ADGRF4, HS3ST5, SEC61G, TSPAN14, SPI1, CTSH, FOXF1, MAPT and WNT3; ADGRF2 and CD2AP were additionally implicated by multi-method or positional mapping at rs10948363. No candidate gene overlapped directly between the female-specific and male-specific sets. However, BIN1 was represented in both the female-specific and shared annotations, whereas CD2AP, MAPT, SPI1 and TSPAN14 occurred in both the male-specific and shared annotations.

Tissue annotations indicated that the identified loci were predominantly represented in central nervous system and immune/peripheral regulatory contexts (**Supplementary Fig. S7**). Among the 127 both-sex/shared regions, eQTL annotations most frequently involved the brain cortex (49 regions), hippocampus (40), frontal cortex BA9 (37), whole blood (32) and spleen (24). All three female-specific regions had cortex eQTL annotations, two were additionally annotated in the hippocampus and frontal cortex, and the TREM2 region was also represented in whole blood and spleen. Among the 14 male-specific regions, cortex, hippocampus and frontal cortex eQTL annotations were observed in eight, seven and six regions, respectively. Whole blood, spleen and EBV-transformed lymphocytes were also recurrently represented in the male-specific set. Chromatin-interaction annotations further highlighted hippocampal and cortical regulatory contexts, together with the caudate, putamen and peripheral immune tissues. Collectively, these annotation patterns suggest that sex-related heterogeneity in AD susceptibility may involve differences in both the genes implicated and the tissue contexts through which female- and male-specific loci exert their effects.

**Supplementary Figures 1. Cohort design, model architecture and evaluation of the STAGE-AD framework.**

Panel **Top**, Discovery analyses included 449,335 participants of European ancestry from ADGC, ADSP, EADB, UK Biobank and All of Us, comprising 259,298 females and 190,037 males. Independent European validation cohorts comprised HUNT and MVP, with 108,446 females and 65,198 males. East Asian validation cohorts comprised Biobank Japan and the National Biobank of Korea, with 30,081 females and 27,442 males. Sex-stratified Alzheimer's disease (AD) GWAS summary statistics and genotype-by-sex interaction results were integrated with linkage disequilibrium reference data, molecular QTLs, chromatin accessibility and contact data, tissue expression and single-cell annotations. Panel **Middle**, STAGE-AD represents variants and regulatory contexts as tokens, incorporating genomic-position information and explicit sex conditioning. Transformer layers combine multi-head self-attention, feed-forward networks, residual connections and normalization. Sex-aware gating and feature-wise linear modulation condition the learned representations on sex context. Cross-attention and gated fusion integrate tissue and cell-type information before prediction through multiple task-specific heads, optimized using a combined loss. Training, validation and test partitions were defined using chromosome-stratified linkage disequilibrium blocks, retaining linked variants and regulatory entities within the same partition. Panel **Bottom**, Representative comparators include logistic regression, least absolute shrinkage and selection operator (LASSO), random forest, XGBoost, multilayer perceptron (MLP) and convolutional neural network (CNN). Evaluation assesses discrimination, candidate ranking, calibration and concordance in independent validation cohorts using AUROC, AUPRC, top-*k* recall and Brier scores. Performance graphics are schematic and do not display observed estimates. AUROC, area under the receiver operating characteristic curve; AUPRC, area under the precision–recall curve; QTL, quantitative trait locus.

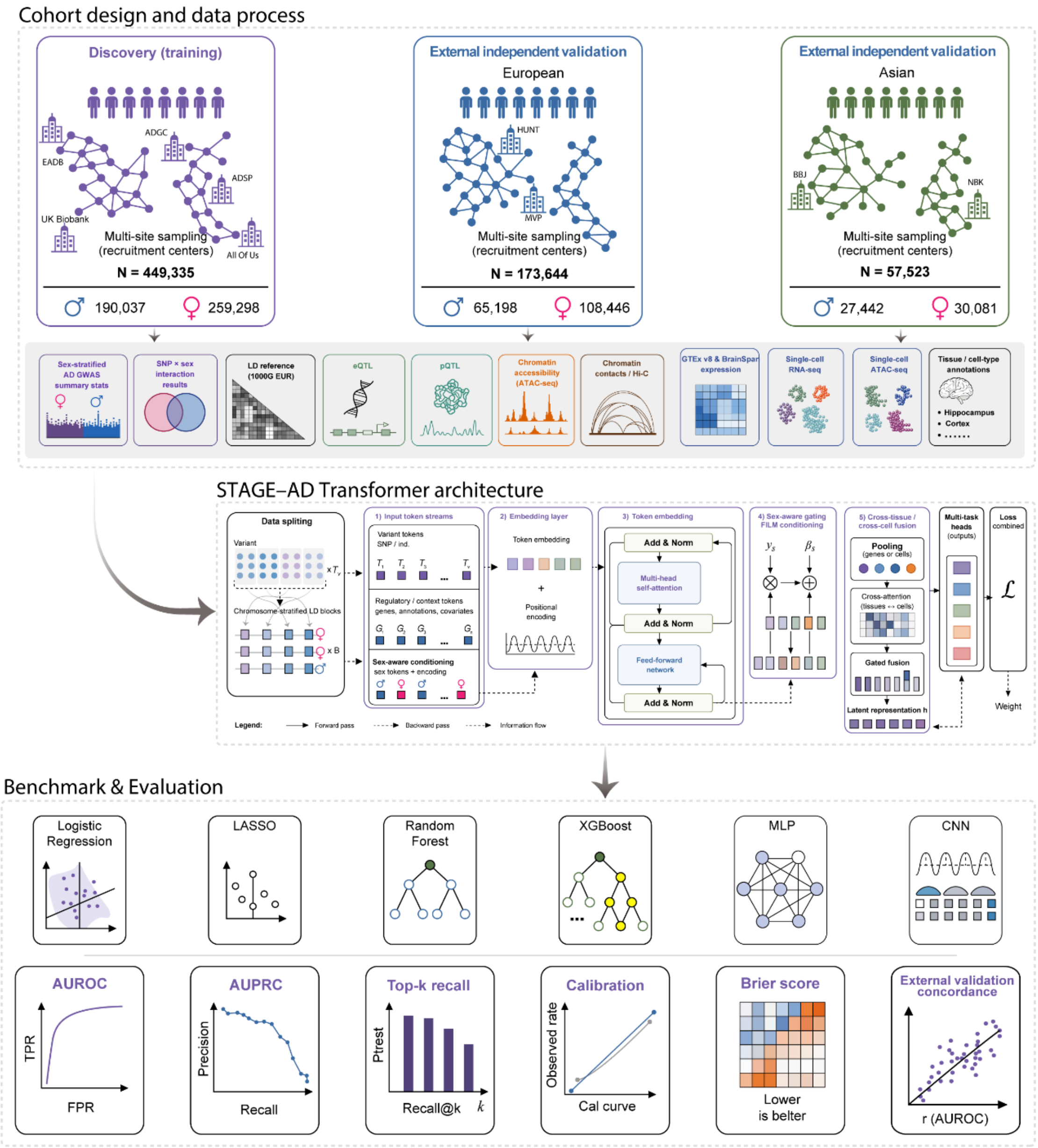

Cohort design and data process
Discovery (training)
EADB
ADGC
ADSP
UK Biobank
All Of Us
Multi-site sampling (recruitment centers)
N = 449,335
190,037
259,298
External independent validation
European
HUNT
MVP
Multi-site sampling (recruitment centers)
N = 173,644
65,198
108,446
External independent validation
Asian
BBJ
NBK
Multi-site sampling (recruitment centers)
N = 57,523
27,442
30,081
Sex-stratified AD GWAS summary stats
SNP × sex interaction results
LD reference (1000G EUR)
eQTL
pQTL
Chromatin accessibility (ATAC-seq)
Chromatin contacts / Hi-C
GTEx v8 & BrainSpan expression
Single-cell RNA-seq
Single-cell ATAC-seq
Tissue / cell-type annotations
• Hippocampus
• Cortex
STAGE–AD Transformer architecture
Data spliting
Chromosome-stratified LD blocks
1) Input token streams
Variant tokens
Regulatory / context tokens
Sex-aware conditioning
2) Embedding layer
Token embedding
Positional encoding
3) Token embedding
Add & Norm
Multi-head self-attention
Feed-forward network
4) Sex-aware gating FiLM conditioning
5) Cross-tissue / cross-cell fusion
Pooling
Cross-attention
Gated fusion
Latent representation h
Multi-task heads (outputs)
Loss combined
Weight
Legend:
Forward pass
Backward pass
Information flow
Benchmark & Evaluation
Logistic Regression
LASSO
Random Forest
XGBoost
MLP
CNN
AUROC
TPR
FPR
AUPRC
Precision
Recall
Top-k recall
Ptrest
Recall@k
Calibration
Observed rate
Cal curve
Brier score
Lower is belter
External validation concordance
r (AUROC)

**Supplementary Figures 2. Genetic correlations (rg) between our female and male genome-wide association studies (GWAS) for Alzheimer's Disease (AD) with previously published GWAS.**

Rg were estimated using linkage disequilibrium score regression (LDSC)[4] using our sex-stratified GWAS summary statistics (Females: 109,525 cases, 149,773 controls. Males: 69,423 cases, 120,614 controls) and publicly available GWAS datasets for AD; the largest published GWAS for AD (EADB et al.) and previous sex-stratified GWAS for AD (Deming et al. and Fan et al.). Points represent the rg point estimates and error bars denote the 95% confidence interval. Females are in pink and males in blue. Stars indicate a significantly different rg between our AD GWAS in females with the previously published AD GWAS versus the rg between our AD GWAS in males with the same previously published AD GWAS. Assessed using the jack-knife method and a two-sided Z-test on the difference in rg across the 200 jack-knife pseudo-values. P-values were adjusted using the Benjamini-Hochberg method for five tests.

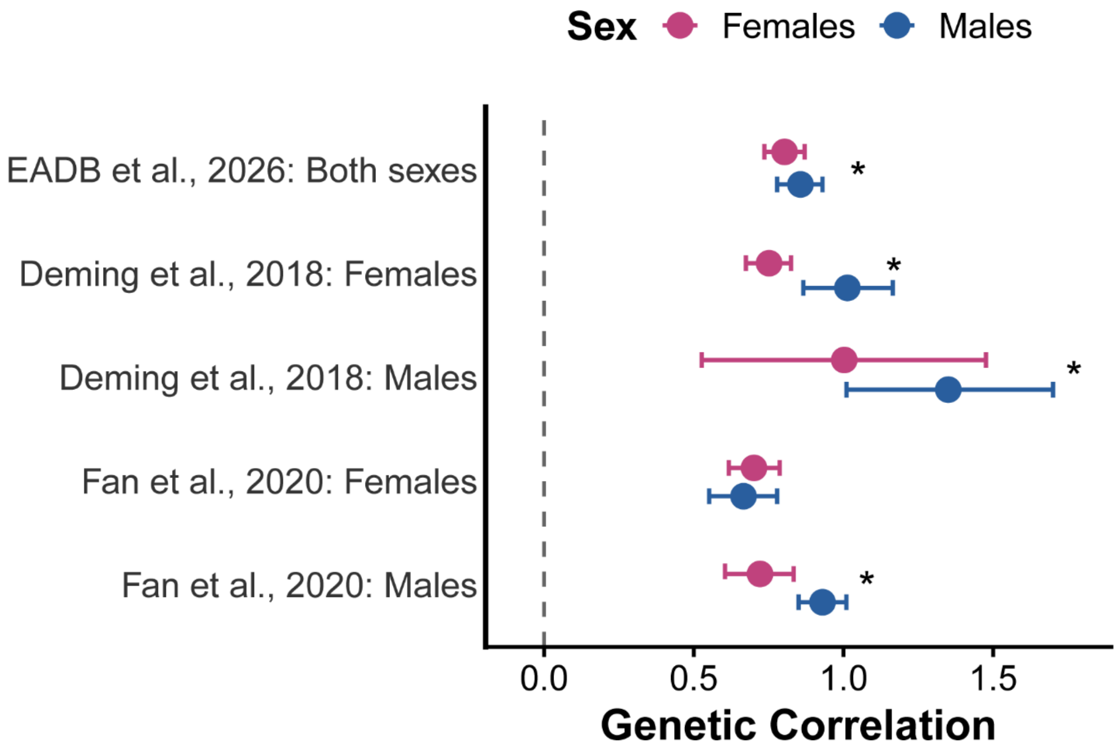

**Supplementary Figures 3. QQ and Miami plots for sex-stratified and genotype-by-sex interaction (G×S) GWAS meta-analysis of Alzheimer's Disease (AD), with female and male meta-analyses shown on the top and bottom, respectively.**

Presented for **a** sex-stratified and **b** G×S-interaction. Lambda value and linkage disequilibrium score regression (LDSC)[5] intercept showed below the QQ plot. The two-sided, unadjusted –log10 p values for GWAS results of each single nucleotide polymorphism (SNP) are shown with positions according to human genome build 38 (GRCh38 assembly). Chromosome 23 is the X chromosome. The darker grey and lighter grey dotted horizontal lines indicate genome-wide significance (P = 5E-08) and suggestive significance (P = 1E-06), respectively. SNPs in dark purple indicate the lead independent genome-wide significant SNPs, and any SNPs in linkage disequilibrium with them. Females: 109,525 cases, 149,773 controls. Males: 69,423 cases, 120,614 controls.

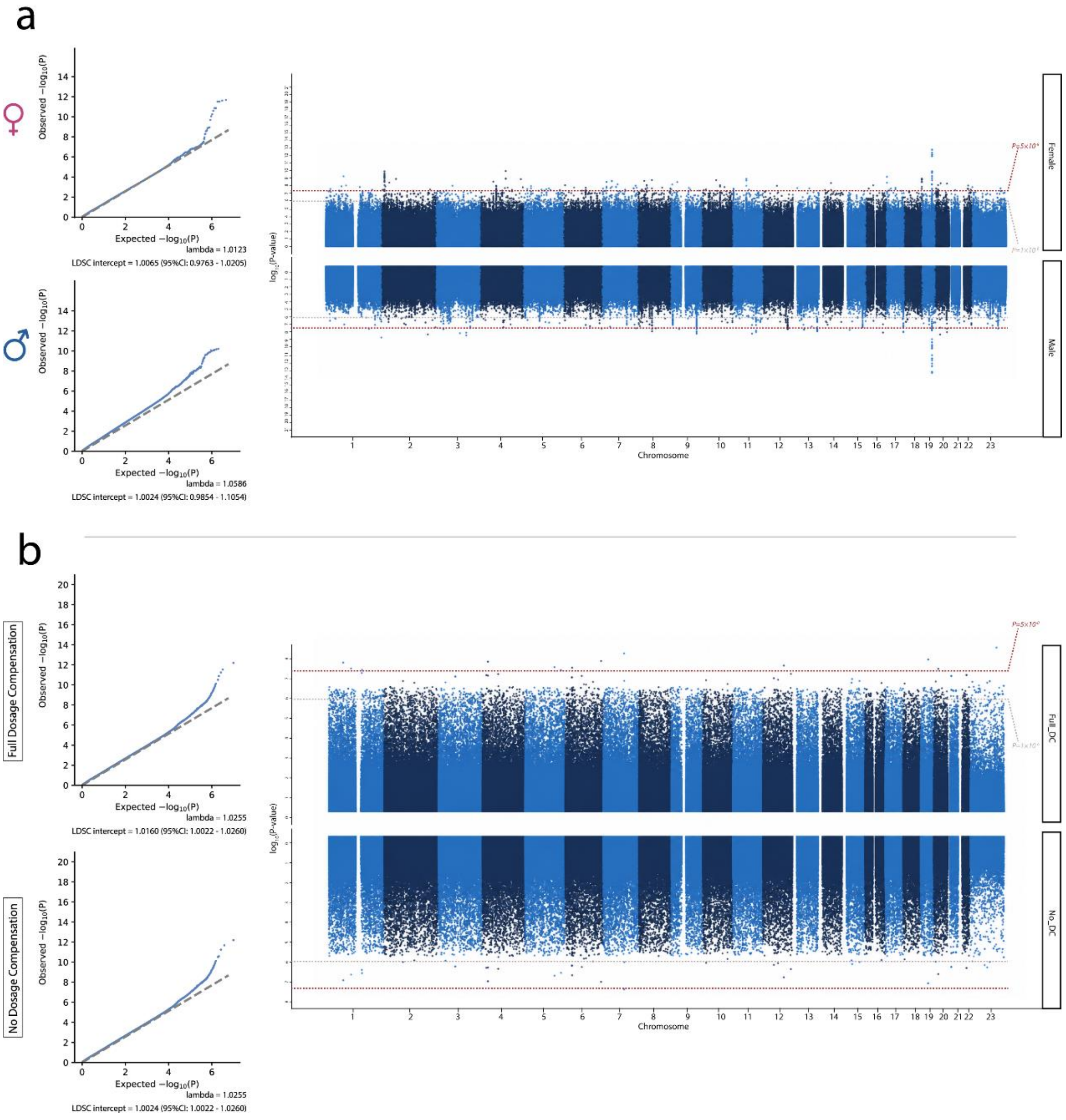

**Supplementary Figures 4. Permutation-based pairwise feature-interaction analysis in the STAGE-AD Transformer and bootstrap AUC distributions across models.**

Panel **a,** Permutation-based pairwise interaction-strength matrices for five feature domains: variant-level features, gene and regulatory-element features, tissue-context features, cell-type-context features and sex-condition features. For each domain, the left and right matrices show interaction scores derived from changes in area under the receiver operating characteristic curve (IAUC) and average precision (IAP), respectively. Interaction strength was evaluated by comparing the performance decrease after jointly permuting a feature pair with the sum of the decreases obtained after permuting each feature separately. Red cells indicate positive, greater-than-additive interactions; blue cells indicate negative interactions; and near-white cells indicate weak or approximately additive effects. The strongest positive interactions were concentrated among variant-level GWAS statistical features, including effect size, standard error, z score and *P* value, whereas molecular-QTL, regulatory, tissue-context and cell-type-context features generally showed weaker pairwise interactions and predominantly complementary marginal contributions. Panel **b,** Density distributions of AUC values obtained from 10,000 bootstrap replicates for the STAGE-AD Transformer (blue), multilayer perceptron (MLP; orange), logistic regression (green), XGBoost (red), convolutional neural network (CNN; purple) and random forest (cyan). Distributions shifted farther to the right indicate better discriminative performance, whereas narrower distributions indicate lower bootstrap variability. The STAGE-AD Transformer showed the most right-shifted and concentrated AUC distribution.

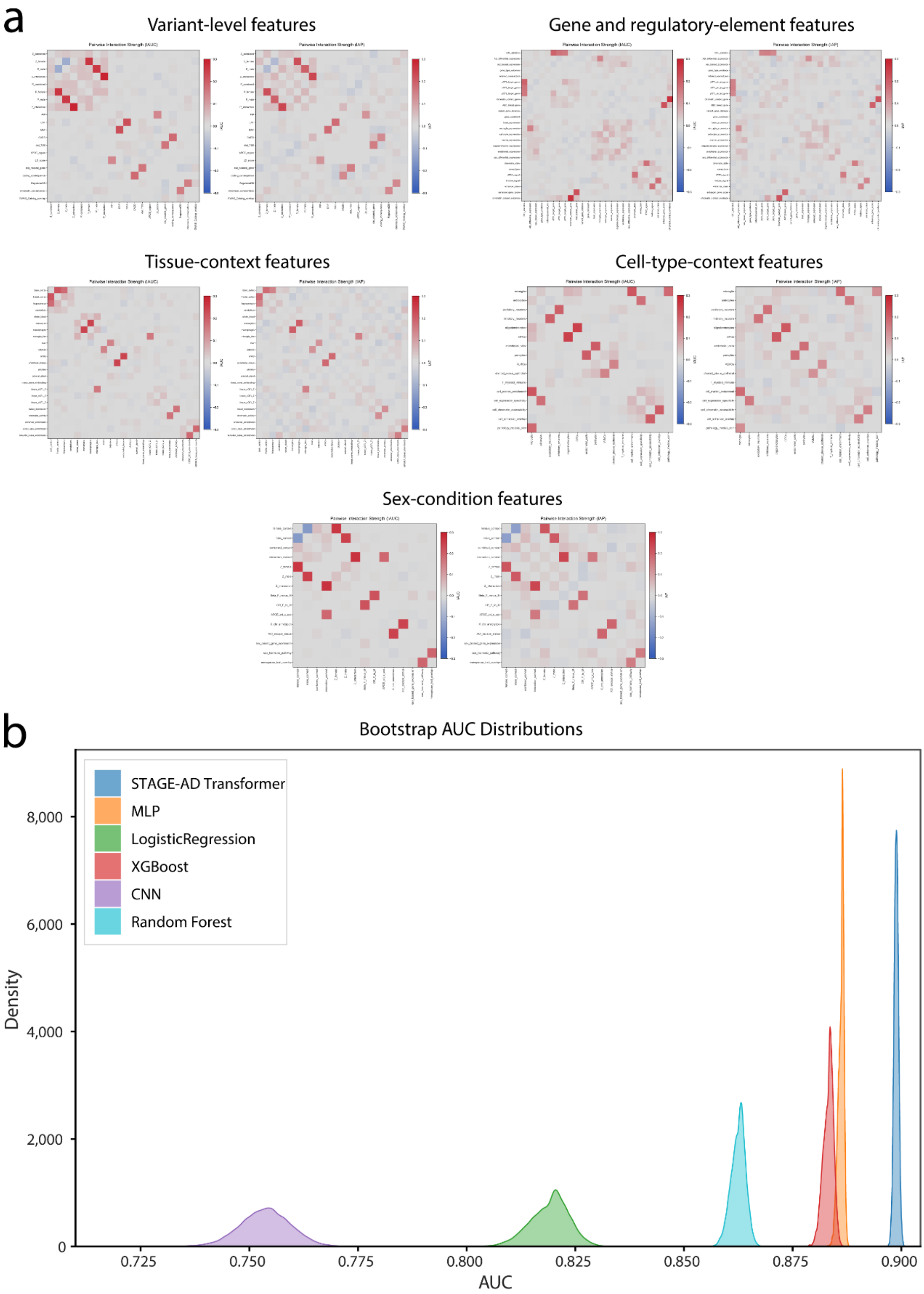
a
Variant-level features
Gene and regulatory-element features
Tissue-context features
Cell-type-context features
Sex-condition features
b
Bootstrap AUC Distributions
STAGE-AD Transformer
MLP
LogisticRegression
XGBoost
CNN
Random Forest
Density
8,000
6,000
4,000
2,000
0
0.725
0.750
0.775
0.800
0.825
0.850
0.875
0.900
AUC

**Supplementary Figures 5. Tissue- and cell-type-ablation analyses of female- and male-biased predictions by STAGE-AD.**

Panel **a**, Leave-one-tissue-out ablation analysis. Bars show the decrease in area under the precision–recall curve (AUPRC) after annotations from each indicated tissue were removed, relative to the full model. Removal of hippocampal annotations produced the largest reduction for female-biased predictions (ΔAUPRC = 0.082), whereas removal of artery–aorta annotations produced the largest reduction for male-biased predictions (ΔAUPRC = 0.074). Panel **b**, Cell-type and cell-state masking analysis. Bars show the decrease in normalized locus-priority score after masking the indicated cell-type or cell-state tokens. Disease-associated/activated microglia produced the largest reduction in female-biased priority scores (Δscore = 0.142), whereas capillary endothelial cells produced the largest reduction in male-biased priority scores (Δscore = 0.129). Male-biased predictions also showed substantial dependence on oligodendrocyte and other endothelial or vascular annotations, whereas female-biased predictions were particularly sensitive to microglial and reactive astrocytic annotations. Female-biased and male-biased results are shown in pink and blue, respectively. Black outlines indicate the largest reduction within each sex category in each panel. Larger decreases indicate greater dependence of model predictions on the corresponding tissue or cellular context. DAM, disease-associated microglia; GFAP, glial fibrillary acidic protein; OPC, oligodendrocyte precursor cell; COP, committed oligodendrocyte precursor; VLMC, vascular leptomeningeal cell.

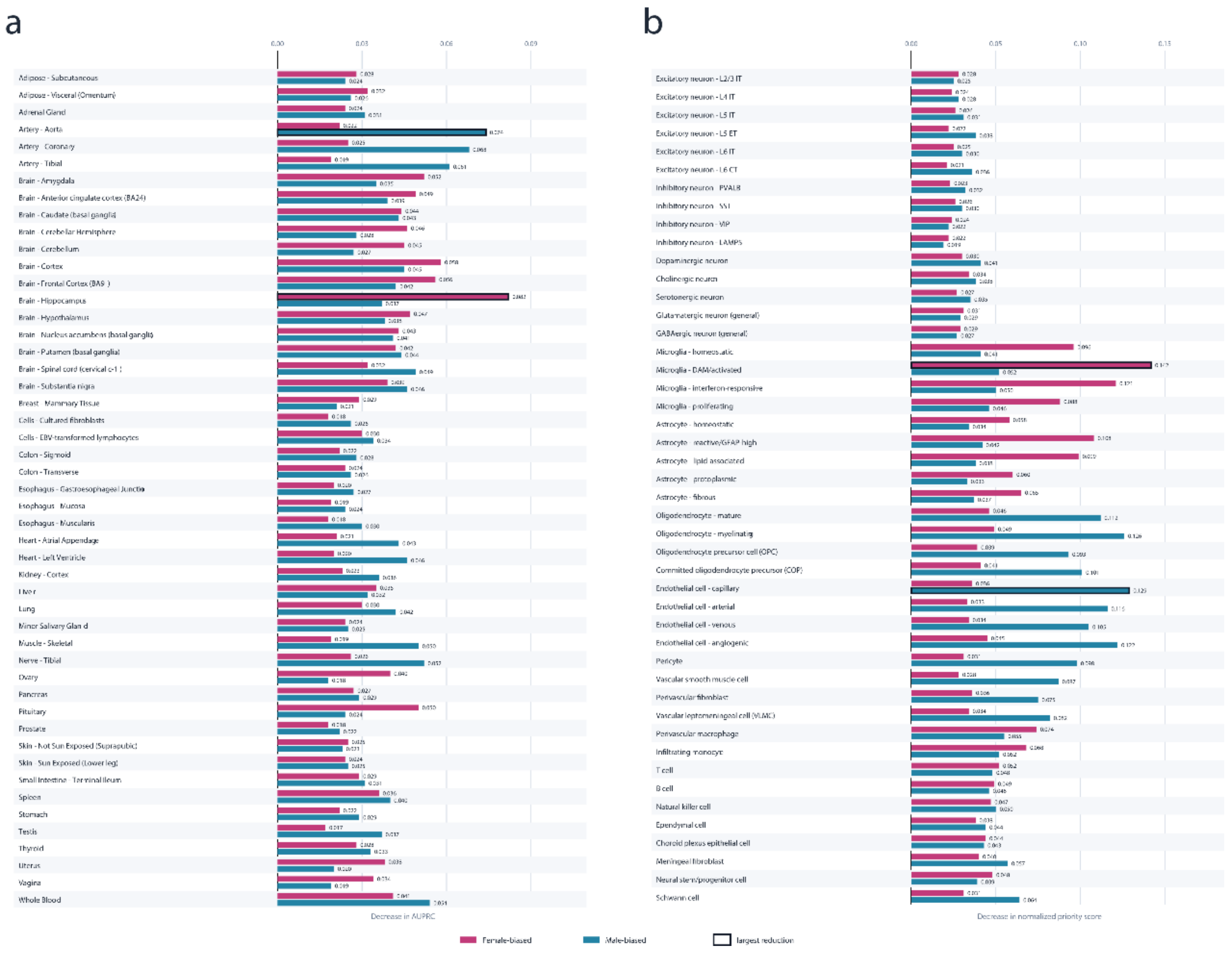
a
0.00
0.03
0.06
0.09
Adipose - Subcutaneous
Adipose - Visceral (Omentum)
Adrenal Gland
Artery - Aorta
Artery - Coronary
Artery - Tibial
Brain - Amygdala
Brain - Anterior cingulate cortex (BA24)
Brain - Caudate (basal ganglia)
Brain - Cerebellar Hemisphere
Brain - Cerebellum
Brain - Cortex
Brain - Frontal Cortex (BA9)
Brain - Hippocampus
Brain - Hypothalamus
Brain - Nucleus accumbens (basal ganglia)
Brain - Putamen (basal ganglia)
Brain - Spinal cord (cervical c-1)
Brain - Substantia nigra
Breast - Mammary Tissue
Cells - Cultured fibroblasts
Cells - EBV-transformed lymphocytes
Colon - Sigmoid
Colon - Transverse
Esophagus - Gastroesophageal Junctio
Esophagus - Mucosa
Esophagus - Muscularis
Heart - Atrial Appendage
Heart - Left Ventricle
Kidney - Cortex
Liver
Lung
Minor Salivary Gland
Muscle - Skeletal
Nerve - Tibial
Ovary
Pancreas
Pituitary
Prostate
Skin - Not Sun Exposed (Suprapubic)
Skin - Sun Exposed (Lower leg)
Small Intestine - Terminal Ileum
Spleen
Stomach
Testis
Thyroid
Uterus
Vagina
Whole Blood
Decrease in AUPRC
b
0.00
0.05
0.10
0.15
Excitatory neuron - L2/3 IT
Excitatory neuron - L4 IT
Excitatory neuron - L5 IT
Excitatory neuron - L5 ET
Excitatory neuron - L6 IT
Excitatory neuron - L6 CT
Inhibitory neuron - PVALB
Inhibitory neuron - SST
Inhibitory neuron - VIP
Inhibitory neuron - LAMP5
Dopaminergic neuron
Cholinergic neuron
Serotonergic neuron
Glutamatergic neuron (general)
GABAergic neuron (general)
Microglia - homeostatic
Microglia - DAM/activated
Microglia - interferon-responsive
Microglia - proliferating
Astrocyte - homeostatic
Astrocyte - reactive/GFAP high
Astrocyte - lipid associated
Astrocyte - protoplasmic
Astrocyte - fibrous
Oligodendrocyte - mature
Oligodendrocyte - myelinating
Oligodendrocyte precursor cell (OPC)
Committed oligodendrocyte precursor (COP)
Endothelial cell - capillary
Endothelial cell - arterial
Endothelial cell - venous
Endothelial cell - angiogenic
Pericyte
Vascular smooth muscle cell
Perivascular fibroblast
Vascular leptomeningeal cell (VLMC)
Perivascular macrophage
Infiltrating monocyte
T cell
B cell
Natural killer cell
Ependymal cell
Choroid plexus epithelial cell
Meningeal fibroblast
Neural stem/progenitor cell
Schwann cell
Decrease in normalized priority score
Female-biased
Male-biased
largest reduction

**Supplementary Figures 6. Sex differences in the genetic architecture and polygenic overlap of Alzheimer's disease.**

Panel **a**–**c**, Posterior distributions of **a**, autosomal SNP-based heritability ($h^2_{SNP}$) on the liability scale, assuming population prevalences of 0.20 in females and 0.10 in males; **b**, polygenicity ($\pi$); and **c**, the selection parameter (S), estimated using SBayesS[6] and sex-stratified GWAS meta-analysis summary statistics. Violin plots show the posterior distributions, points indicate posterior means, and vertical lines indicate 95% highest posterior density intervals. The posterior probability that the female estimate exceeds the male estimate, P(F>M), is shown above each panel. Panel **d**, Autosomal SNP-based genetic correlations (rg) between female and male AD estimated using linkage disequilibrium score regression. Results are shown for the sex-stratified meta-analysis summary statistics and for six within-cohort female–male, 30 across-cohort female–male, 15 across-cohort male–male and 15 across-cohort female–female comparisons. Panel **e**, Pearson correlations between standardized female and male effect-size estimates for lead independent genome-wide significant SNPs identified in the largest sex-combined AD GWAS, presented using the same cohort-comparison groups as in panel **d**. In panel **d** and **e**, pale points represent estimates from individual comparisons, large points represent pooled estimates, and horizontal lines indicate 95% confidence intervals. The vertical dashed line denotes a correlation of 1. Asterisks indicate correlations significantly different from 1 using two-sided Z tests, with Benjamini–Hochberg adjustment across five comparisons. Female–female, male–male and female–male comparisons are shown in pink, blue and orange, respectively. Panel **f**, Bivariate MiXeR[7] estimates of the numbers and proportions of causal variants explaining 90% of ($h^2_{SNP}$): 219 (1.24%) female-only variants, 17,266 (97.98%) variants shared between females and males and 137 (0.78%) male-only variants. The estimated correlation of effect sizes among shared causal variants is displayed below the diagram. Panel **g**, gwas-pw[8] classification of 127 autosomal genomic regions with posterior probability of association greater than 0.5: 3 (2.36%) female-specific regions, 120 (94.49%) regions shared between sexes and 4 (3.15%) male-specific regions. Female-specific and male-specific components are shown in pink and blue, respectively. Analyses included 109,525 female cases and 149,773 female controls, and 69,423 male cases and 120,614 male controls.

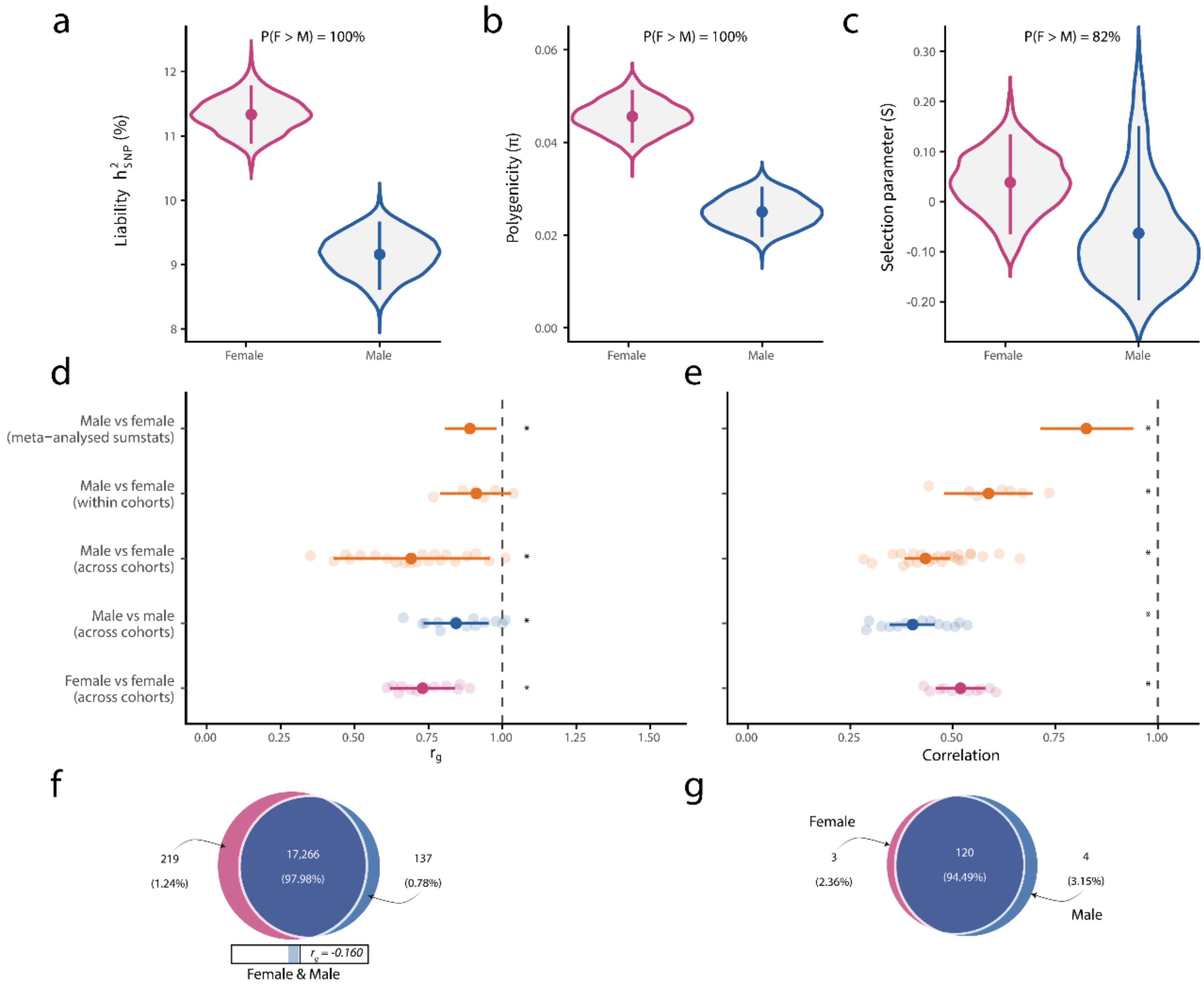
a
P(F > M) = 100%
Liability $h^2_{SNP}$ (%)
Female
Male
b
P(F > M) = 100%
Polygenicity (π)
c
P(F > M) = 82%
Selection parameter (S)
d
Male vs female (meta−analysed sumstats)
Male vs female (within cohorts)
Male vs female (across cohorts)
Male vs male (across cohorts)
Female vs female (across cohorts)
$r_g$
e
Correlation
f
219
(1.24%)
17,266
(97.98%)
137
(0.78%)
$r_g$ = -0.160
Female & Male
g
Female
3
(2.36%)
120
(94.49%)
4
(3.15%)
Male

**Supplementary Figures 7. Miami plot of sex-stratified genome-wide association study (GWAS) meta-analysis of Alzheimer's Disease (AD), with genomic regions identified by gwas-pw to contain causal risk loci for AD that are shared between females and males or are sex-specific.**

Female and male GWAS meta-analysis results are displayed in the upper and lower panels, respectively, with the male results mirrored for visualization. Each point represents the two-sided, unadjusted $-\log_{10}$ P value for one single-nucleotide polymorphism (SNP), plotted according to its genomic position in the GRCh38 assembly. Alternating light- and dark-blue points distinguish adjacent chromosomes. Chromosome 23 denotes the X chromosome; gwas-pw analysis was restricted to autosomal regions. The red and light-grey dotted horizontal lines indicate the genome-wide significance threshold (P = 5E-08) and suggestive association threshold (P = 1E-06), respectively. Vertical bands indicate 127 autosomal regions with a gwas-pw posterior probability of association greater than 0.5: grey bands denote 120 regions inferred to contain a causal risk signal shared between females and males, pink bands denote three female-specific regions, and dark blue bands denote four male-specific regions. Analyses included 109,525 female cases and 149,773 female controls, and 69,423 male cases and 120,614 male controls.

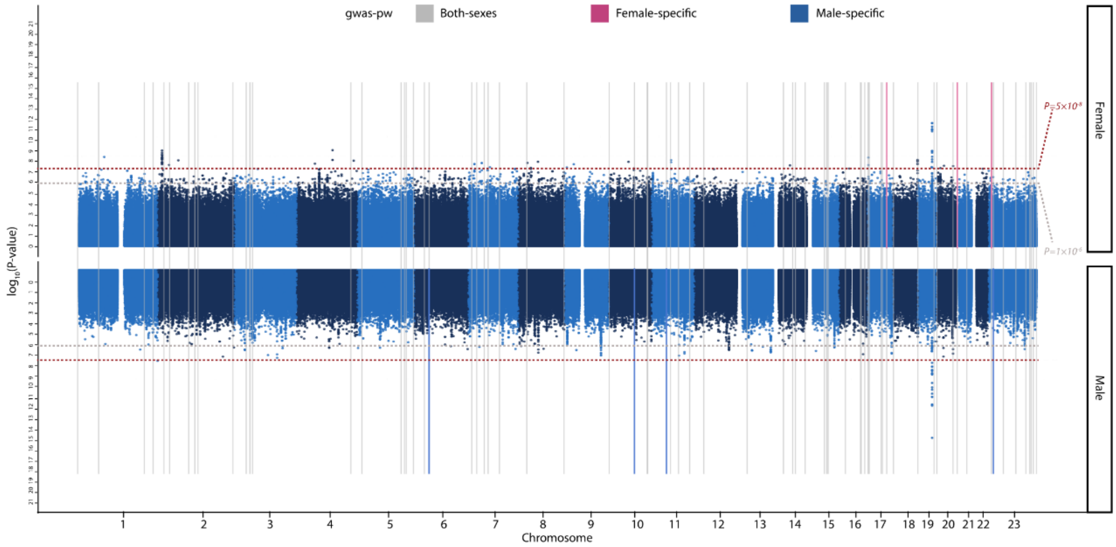

**Supplementary Figures 8. FUMA gene-property analyses of tissue-specific gene expression for sex-stratified Alzheimer's Disease (AD) GWAS.**

FUMA gene-property analyses evaluated whether genes with higher expression in particular developmental stages or tissues showed stronger gene-level associations with AD in the female and male GWAS meta-analyses. Panel **a**, Results for 11 general developmental stages of human brain samples from BrainSpan. Panel **b**, Results for 30 general tissue categories from GTEx v8[9]. Panel **c**, Results for 53 specific tissues from GTEx v8. Bars represent the $-\log_{10}$-transformed unadjusted P values, with female and male results shown in pink and blue, respectively. Grey dashed horizontal lines indicate the corresponding Bonferroni-corrected significance thresholds: P = 0.05/11 in **a**, P = 0.05/30 in **b**, and P = 0.05/53 in **c**. No developmental stage or tissue exceeded its panel-specific Bonferroni threshold. Females are in pink and males in dark blue. Females: 109,525 cases, 149,773 controls. Males: 69,423 cases, 120,614 controls.

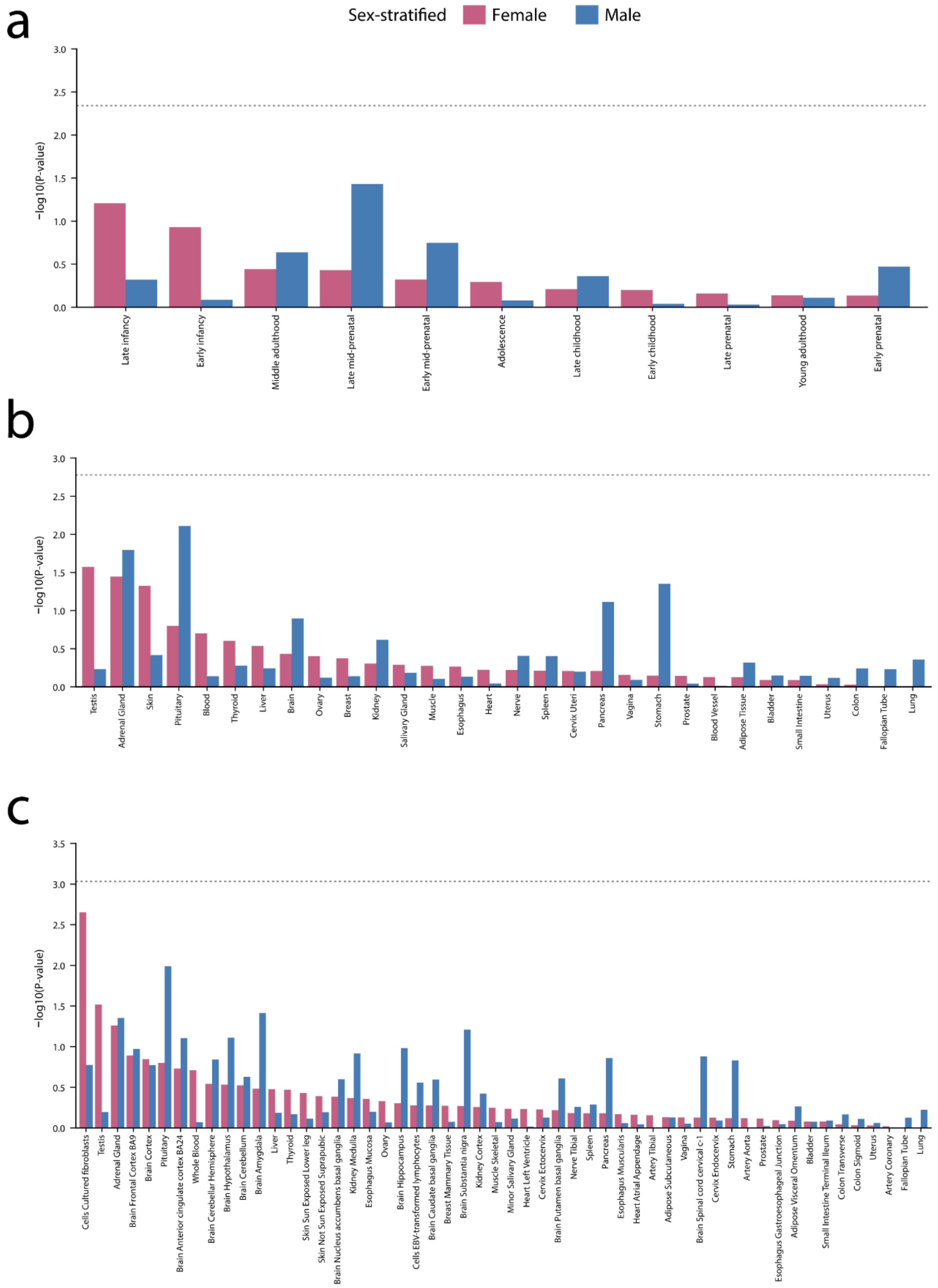
a
Sex-stratified
Female
Male
−log10(P-value)
Late infancy
Early infancy
Middle adulthood
Late mid-prenatal
Early mid-prenatal
Adolescence
Late childhood
Early childhood
Late prenatal
Young adulthood
Early prenatal
b
−log10(P-value)
Testis
Adrenal Gland
Skin
Pituitary
Blood
Thyroid
Liver
Brain
Ovary
Breast
Kidney
Salivary Gland
Muscle
Esophagus
Heart
Nerve
Spleen
Cervix Uteri
Pancreas
Vagina
Stomach
Prostate
Blood Vessel
Adipose Tissue
Bladder
Small Intestine
Uterus
Colon
Fallopian Tube
Lung
c
−log10(P-value)
Cells Cultured fibroblasts
Testis
Adrenal Gland
Brain Frontal Cortex BA9
Brain Cortex
Pituitary
Brain Anterior cingulate cortex BA24
Whole Blood
Brain Cerebellar Hemisphere
Brain Hypothalamus
Brain Cerebellum
Brain Amygdala
Liver
Thyroid
Skin Sun Exposed Lower leg
Skin Not Sun Exposed Suprapubic
Brain Nucleus accumbens basal ganglia
Kidney Medulla
Esophagus Mucosa
Ovary
Brain Hippocampus
Cells EBV-transformed lymphocytes
Brain Caudate basal ganglia
Breast Mammary Tissue
Brain Substantia nigra
Kidney Cortex
Muscle Skeletal
Minor Salivary Gland
Heart Left Ventricle
Cervix Ectocervix
Brain Putamen basal ganglia
Nerve Tibial
Spleen
Pancreas
Esophagus Muscularis
Heart Atrial Appendage
Artery Tibial
Adipose Subcutaneous
Vagina
Brain Spinal cord cervical c-1
Cervix Endocervix
Stomach
Artery Aorta
Prostate
Esophagus Gastroesophageal Junction
Adipose Visceral Omentum
Bladder
Small Intestine Terminal Ileum
Colon Transverse
Colon Sigmoid
Uterus
Artery Coronary
Fallopian Tube
Lung

**Supplementary Figures 9. Illustrative brain cell-type distributions and candidate regulatory contacts.**

Panel **a-b**, UMAP-style schematics depicting 21 annotated neuronal, glial, vascular, epithelial and immune cell populations. Colors denote cell types, with matching colors used across panels. OPCs, oligodendrocyte precursor cells; VLMCs, vascular leptomeningeal cells. Panel **c-d**, Chromatin-contact maps centered on the candidate regulatory elements GH13J023887 (chr13: 23,889,180–23,889,680; **c**) and GH04J055126 (chr4: 55,126,321–55,126,987; **d**). Red intensity indicates normalized interaction frequency, with darker colors denoting higher contact frequencies. Black boxes and red vertical marks indicate the highlighted regulatory elements. Blue gene tracks show neighboring genes and their transcriptional orientations. Tables summarize the reported promoter contacts, signed genomic distances, interaction frequencies and q values. The displayed candidate target genes are MIPEP, PCOTH, C1QTNF9B, TNFRSF19 and C1QTNF9 in **c**, and KDR, SRD5A3 and PDGFRA in **d**.

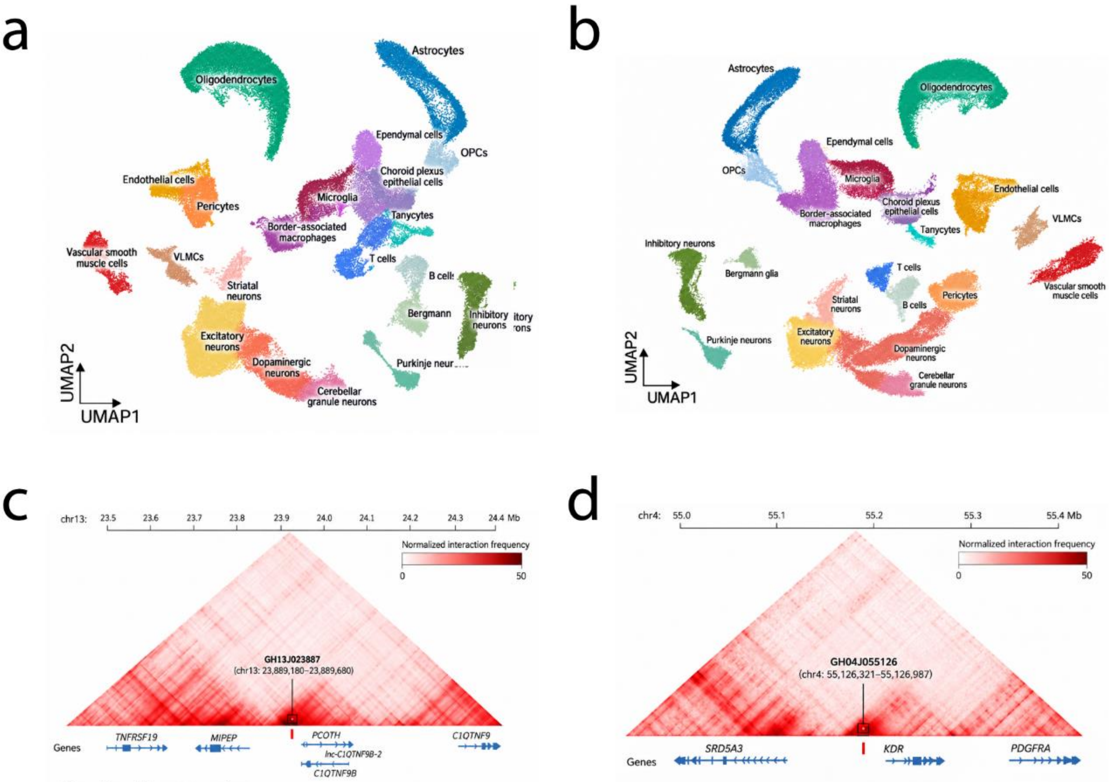


Interaction with gene promoters

| Target gene | Distance (kb) | Interaction frequency | *q* value |
|---|---|---|---|
| *MIPEP* | −156 | 22.8 | $3.4 \times 10^{-6}$ |
| *PCOTH* | +28 | 18.1 | $2.7 \times 10^{-5}$ |
| *C1QTNF9B* | +112 | 12.6 | $6.1 \times 10^{-5}$ |
| *TNFRSF19* | −315 | 8.4 | $1.9 \times 10^{-4}$ |
| *C1QTNF9* | +422 | 6.3 | $8.7 \times 10^{-4}$ |

Interaction with gene promoters

| Target gene | Distance (kb) | Interaction frequency | *q* value |
|---|---|---|---|
| *KDR* | +72 | 18.4 | $2.1 \times 10^{-6}$ |
| *SRD5A3* | −118 | 7.6 | $4.3 \times 10^{-4}$ |
| *PDGFRA* | +310 | 5.2 | $1.7 \times 10^{-3}$ |

**Supplementary Figures 10. Drug-signature mapping across sex-aware Alzheimer's Disease (AD) regulatory modules.**

Sex-aware AD expression signatures were constructed from expression-based transcriptome-wide association study gene-level Z-scores and compared with drug-induced expression profiles from the LINCS Connectivity Map Level 5 replicate-consensus dataset[10]. Curves show the tested drug signatures ranked by their ensemble connectivity scores. Negative scores indicate compounds predicted to reverse the corresponding AD-associated expression signature, whereas positive scores indicate concordant transcriptional effects; dashed horizontal lines denote a score of zero. Red circles and grey callouts identify representative compounds from the negative tail of each distribution: panel **a**, lenalidomide for the female-biased module; panel **b**, mycophenolic acid for the male-biased module; panel **c**, sirolimus for the shared module; and panel **d**, lenalidomide for the genotype-by-sex interaction module. Inset heatmaps compare the drug-induced and AD-associated expression signatures across selected genes. Purple and teal indicate positive and negative gene-expression Z-scores, respectively; opposing color patterns between the drug and disease signatures indicate predicted transcriptional reversal. These rankings represent hypothesis-generating drug-repurposing signals and do not establish therapeutic efficacy, clinical safety or a beneficial direction of treatment effect.

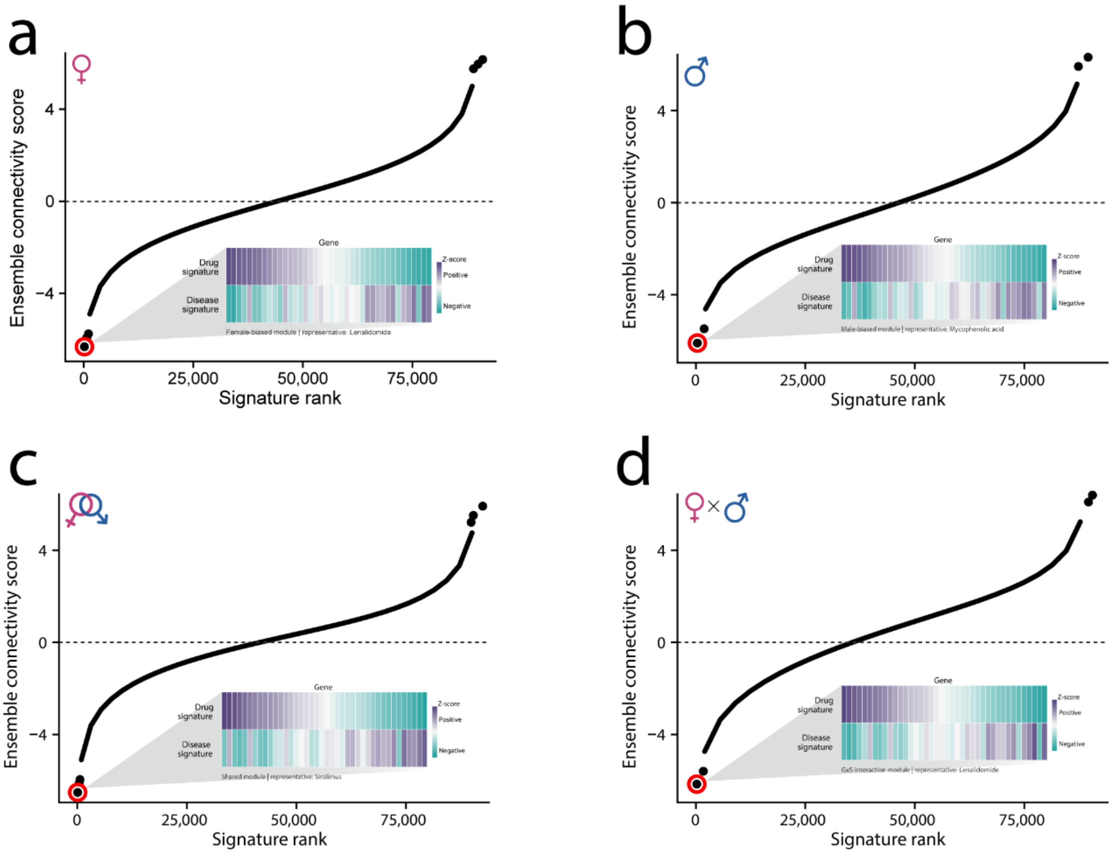

**Supplementary Methods 1**

**Discovery and external validation cohorts**

The discovery analyses included the Alzheimer's Disease Genetics Consortium (ADGC), Alzheimer's Disease Sequencing Project (ADSP), European Alzheimer & Dementia Biobank (EADB), All of Us Research Program and UK Biobank. External validation used the Trøndelag Health Study (HUNT), Million Veteran Program (MVP), BioBank Japan (BBJ) and National Biobank of Korea (NBK). Cohort-specific phenotype definitions, genomic processing and association procedures are described below. Contributing studies obtained institutional review board or ethics committee approval, and participants or their legally authorized representatives provided informed consent.

**Shared association procedures**

For ADGC, ADSP, All of Us, HUNT, MVP, BBJ and NBK, autosomal single-nucleotide polymorphisms (SNPs) were filtered and linkage disequilibrium (LD)-pruned in PLINK v1.9[11] using --maf 0.01, --geno 0.02, --mind 0.02, --hwe 1e-10 and --indep-pairwise 1500 100 0.2. These variants were used to construct a genetic relationship matrix (GRM) and a sparse GRM with a relatedness threshold of 0.03 in GCTA v1.94.1. GRMs were constructed within the ancestry group analyzed: European ancestry for ADGC, ADSP, All of Us, HUNT and MVP; Japanese ancestry for BBJ; and Korean or East Asian ancestry for NBK. Association analyses used fastGWA[12] in GCTA v1.94.1, with the first ten ancestry principal components (PCs) included in sex-stratified and genotype-by-sex (G×S) analyses. Additional cohort-specific covariates and departures from these procedures are specified below.

**Discovery cohorts**

**Alzheimer's Disease Genetics Consortium**

Participants were recruited primarily through US Alzheimer's Disease Research Centers and associated studies.[13] Alzheimer's disease (AD) cases had a clinical diagnosis based on standardized cognitive and neurological assessments or neuropathological confirmation based on amyloid plaques and neurofibrillary tangles. Controls had no dementia, mild cognitive impairment (MCI) or other relevant neurodegenerative condition at assessment; autopsied controls had low or absent AD neuropathology. Exclusions included major non-AD neurological disorders, non-AD tauopathies, synucleinopathies, Down syndrome and other conditions that could confound diagnosis, as specified by the contributing studies.

Participants were genotyped using the Illumina Global Screening Array. Genetic ancestry was assessed with smartpca (build 1000) using approximately 37,000 thinned SNPs and PC axes defined by 1000 Genomes Project reference individuals. Participants more than six standard deviations from the European-reference centroid on PC1 or PC2 were excluded. Pre-imputation variant quality control used GenomeStudio v2.0 and PLINK v1.9; variants were excluded for a GenTrain score <0.6, minor allele frequency (MAF) <0.01, call rate <95% or Hardy–Weinberg equilibrium (HWE) $P < 1 \times 10^{-6}$. HWE testing of X-chromosome variants was restricted to females. Genotypes were imputed against the TOPMed R2 reference panel.[14] Individuals represented in overlapping datasets were removed to limit duplication in meta-analysis. Association testing followed the shared procedures above.

**Alzheimer's Disease Sequencing Project**

Whole-genome sequencing (WGS) data were obtained from ADSP Release 4 (NIAGADS accession NG00067).[15] Participants required harmonized AD case–control status, genetic ancestry information and complete covariates. Cases included definite, probable, possible, prevalent or incident AD according to the ADSP phenotype files; controls were cognitively normal individuals without AD or another dementia. Participants with MCI, non-AD dementia, uncertain diagnosis or missing phenotype information were excluded. Age at onset or diagnosis was used for cases and age at the last cognitive assessment or death for controls. The analytical sample comprised 22,200 cases and 33,222 controls.

WGS was performed across sequencing centers using Illumina platforms, predominantly with PCR-free libraries and 150-bp paired-end reads. Reads were processed using the ADSP Variant Calling Pipeline and data management tool (VCPA), aligned to GRCh38 and jointly genotyped. We used ADSP-provided, quality-controlled biallelic single-nucleotide variant and short insertion/deletion files; no imputation was performed. Low-quality genotype calls were set to missing using the ADSP genotype-level filters. Sample quality control assessed WGS–array genotype concordance, reported and genetically inferred sex, contamination, duplicates and relatedness. Samples with unresolved discordance, contamination or sequencing-quality failures were excluded. VerifyBamID FREEMIX >0.05 indicated possible contamination, and pairs with identity-by-descent PI_HAT >0.4 were reviewed for known or unexpected relationships.

Variants were required to pass ADSP variant-level filters. For population-structure and GRM analyses, the study selected common biallelic variants with an ADSP variant flag of zero, high call rate, HWE *P* above the prespecified threshold and MAF >0.05; long-range LD regions were excluded before pruning. The ADSP Release 4 reference procedure used MAF >0.02, call rate >99.5%, HWE $P > 5 \times 10^{-4}$ and LD $r^2 < 0.05$. Population structure and relatedness were evaluated using PC-AiR and GENESIS, with ancestry additionally assessed against 1000 Genomes reference populations. Analyses were conducted within genetically inferred ancestry groups supported by the available sample size. Association testing of the European-ancestry subset followed the shared procedures above.

**European Alzheimer & Dementia Biobank**

WGS data were obtained through the Alzheimer Disease European Sequencing component of EADB,[16] which includes memory-clinic, research-cohort and population-based participants from multiple European countries. Cases met possible or probable AD criteria under the National Institute on Aging–Alzheimer's Association or NINCDS–ADRDA frameworks, with biomarker or neuropathological support when available. Controls were cognitively normal and had no dementia diagnosis. MCI, non-AD dementia, uncertain diagnosis and missing phenotype information were exclusion criteria. Age at onset or diagnosis was used for cases and age at the latest assessment or death for controls. The analytical sample comprised 20,712 AD cases and 22,445 controls.

Paired-end Illumina reads were processed through a harmonized pipeline, aligned to GRCh38 and jointly called using GATK; no genotype imputation was performed. Sample filters addressed sequencing quality, contamination, sex discordance, duplication, missingness and heterozygosity. Variant filters addressed sequencing quality, read depth, genotype quality, call rate, HWE and minor allele count. Common LD-pruned autosomal variants were used to estimate ancestry, PCs and genetic relatedness. Association analyses used fastGWA in GCTA v1.94.1 and a sparse GRM threshold of 0.05. Pooled models included age, sex, sequencing center or batch, study cohort and the first ten ancestry PCs; sex-stratified models included the same covariates except sex. G×S analyses included genotype and sex main effects and their interaction. X-chromosome pseudoautosomal and non-pseudoautosomal regions were analyzed separately, with hemizygous male non-pseudoautosomal genotypes and sex-specific call-rate, allele-count and HWE filters. Genome-wide significance was defined as $P < 5 \times 10^{-8}$.

**All of Us Research Program**

All of Us integrates genomic data, electronic health records (EHRs), surveys and physical measurements from participants recruited across the United States through healthcare organizations and direct-volunteer pathways.[17] EHRs were harmonized to the Observational Medical Outcomes Partnership Common Data Model. AD cases had ICD-9-CM code 331.0 or ICD-10-CM codes G30.* or F00.*; the earliest qualifying record defined age at diagnosis. Controls were aged ≥65 years at their latest healthcare encounter and had no recorded AD, MCI, dementia or other major neurodegenerative disorder. Participants with uncertain phenotype status or missing covariates were excluded. Analyses were restricted to WGS participants genetically similar to the European reference population (Data repository release R2019Q4R3].

PCR-free WGS was performed on Illumina NovaSeq 6000 instruments at a mean coverage of at least 30×. Reads were processed with Illumina DRAGEN, aligned to GRCh38dh and jointly called; no imputation was performed. Samples and variants were required to pass All of Us quality control, including coverage, contamination, mapping quality, sample identity, genotype concordance, sex concordance, call rate, MAF and HWE criteria. X-chromosome HWE testing was restricted to females. Ancestry was assessed against 1000 Genomes and Human Genome Diversity Project reference individuals using LD-pruned common variants. Association analyses followed the shared procedures, additionally adjusting for age and sequencing batch. G×S models included genotype and sex main effects and their interaction. X-chromosome pseudoautosomal and non-pseudoautosomal regions were processed separately, and male non-pseudoautosomal genotypes were treated as hemizygous.

**UK Biobank**

UK Biobank recruited participants aged 40–69 years across the United Kingdom during 2006–2010 and included 488,377 genotyped participants.[18] AD status was defined using the algorithmic outcome in field 42020 (date of AD report), with ascertainment source from field 42021. This outcome captures prevalent and incident AD using hospital inpatient records and national death registries, including primary or secondary diagnoses and underlying or contributory causes of death (Application number 825570).

For record-based phenotype reconstruction, qualifying codes were ICD-9 331.0 and ICD-10 F00, F00.0, F00.1, F00.2, F00.9, G30, G30.0, G30.1, G30.8, G30.9 and G31.0. Hospital diagnoses were obtained from fields 41202 and 41204 and causes of death from fields 40001 and 40002. The earliest qualifying date distinguished prevalent from incident cases. Self-reported code 1263 in field 20002 was insufficient to define AD because it combines dementia, AD and cognitive impairment, but was used to exclude potentially affected controls. Controls had no algorithmic AD or all-cause dementia outcome (fields 42020 and 42018), no dementia diagnosis in linked records and no self-reported code 1263. Participants with vascular, frontotemporal, unspecified or other dementia subtypes were excluded from both groups, and controls remained free of recorded dementia throughout available follow-up.

A validation study reported an approximately 71% positive predictive value for AD identified from combined healthcare sources.[19] Participants who withdrew consent, lacked required phenotype or covariate information, or were outside the UK Biobank genetically defined White British subset were excluded. Ethical approval was provided by the North West Multi-centre Research Ethics Committee (11/NW/0382).

Centrally quality-controlled, phased and imputed genotypes were used [15]. Autosomal variants were filtered and LD-pruned in PLINK v1.9 using --maf 0.01, --geno 0.02, --mind 0.10, --hwe 1e-10 and --indep-pairwise 1500 150 0.2. These variants were used to construct the GRM and a sparse GRM with a threshold of 0.05 in GCTA v1.94.1, including for analyses involving chromosome X. Imputed variants were required to meet the prespecified MAF and missingness criteria and INFO ≥0.8.

Sex-stratified analyses used fastGWA-GLMM in GCTA v1.94.1 to account for relatedness, population structure and case–control imbalance through a generalized linear mixed model with saddle-point approximation.[20] Covariates were age, age squared, genotyping array, assessment center and the first ten ancestry PCs. Combined-sample G×S models additionally included sex, genotype and their interaction, testing whether the allelic AD log-odds ratio differed between females and males. X-chromosome pseudoautosomal and non-pseudoautosomal regions were analyzed separately; sex information, male genotype coding and dosage-compensation assumptions were checked before testing. X-chromosome analyses used model parameters estimated from autosomal variants and the autosomal sparse GRM.

**External validation cohorts**

**Trøndelag Health Study**

HUNT participants were drawn from HUNT2 (1995–1997) and HUNT3 (2006–2008) in Trøndelag County, Norway.[21] AD and dementia phenotypes were ascertained through regional inpatient and outpatient registries covering 1987–2018, using prespecified ICD-9 and ICD-10 codes. A subset of registry diagnoses had been reviewed by specialists in geriatrics and old-age psychiatry. Prevalent AD was defined by a qualifying diagnosis before or at HUNT participation; incident AD required no recorded dementia at baseline and a qualifying AD diagnosis during follow-up through 2018. Controls had no recorded AD or other dementia during follow-up. Participants with unusable genotypes, missing essential phenotypes or covariates, or failed genetic quality control were excluded. The analytical sample comprised 23,124 AD cases and 38,166 controls, including 39,201 females and 22,089 males (Project number 2020/0081).

DNA from 71,860 participants was genotyped using HumanCoreExome12 v1.0, HumanCoreExome12 v1.1 or the customized UM HUNT Biobank v1.0 array. Genotypes were called in GenomeStudio. Sample exclusions included call rate <99%, contamination >2.5%, large chromosomal copy-number abnormalities, discordance between inferred and reported sex, sex-chromosome constitutions other than XX or XY, or lower call rate within a technical-duplicate or twin pair. Variant exclusions included non-unique or imperfect probe alignment to GRCh37, cluster separation <0.30, GenTrain score <0.15, call rate <99% or HWE $P < 1 \times 10^{-4}$ in unrelated European-ancestry participants. For duplicate assays, the assay with the highest call rate was retained.

Ancestry was assessed by projection onto Human Genome Diversity Project PCs, retaining participants within the European-reference space. Array datasets were harmonized using shared markers. Variants with between-array allele-frequency differences >15%, or monomorphic on one array but with MAF >1% on another, were excluded. Genotypes were phased with Eagle2 v2.3 and imputed in 61,290 participants using Minimac3 v2.0.1. The autosomal reference combined Haplotype Reference Consortium (HRC) release 1.1 with WGS data from 2,201 HUNT participants sequenced at approximately 5×; these sequences served as an imputation reference rather than the primary dataset. Chromosome X was imputed against HRC v1.1. Variants with imputation Rsq <0.30 or minor allele count <3 were excluded. Association testing followed the shared procedures.

**Million Veteran Program**

MVP is a US Department of Veterans Affairs (VA) cohort linking blood samples, questionnaires and EHRs from veterans recruited nationwide.[22] Participants with WGS and AD-related phenotype information were included. AD was ascertained from structured diagnostic codes, supplemented by medication records, clinical encounters and phenotype algorithms. At least one AD-related diagnosis after enrolment defined a case; participants with no recorded AD or related dementia during follow-up were controls. Exclusions included unavailable or low-quality genomic data, discordant sex assignment and missing key covariates. Age and sex were obtained from baseline data, and ancestry was assessed using genetic PCs and clustering. The study was approved by the VA Central Institutional Review Board (Application number 23110578).

Peripheral-blood DNA underwent WGS through the MVP Genomic Analysis Program using paired-end Illumina NovaSeq sequencing, alignment to GRCh38 and standardized variant calling. Sample quality control assessed sequencing quality, contamination, sex concordance, heterozygosity, relatedness and phenotype completeness. Variants were filtered for genotype quality, read depth, call rate and HWE. Genomic location and predicted molecular consequences were annotated, and ancestry was evaluated using common autosomal variants and reference populations. For analyses using imputed dosages, a WGS-based reference framework or imputation panel was used, with filtering for imputation quality and allele count. Association testing of European-ancestry participants followed the shared procedures.

**BioBank Japan**

BBJ is a hospital-based biobank of Japanese patients recruited through collaborating medical institutions.[23] AD cases had a clinical diagnosis in the harmonized phenotype data, supported by standardized assessments and probable or possible AD criteria when detailed records were available. Controls were aged ≥65 years with no recorded AD, MCI, dementia or major neurodegenerative disorder. Participants with uncertain phenotype status or missing age, sex or covariates were excluded. Analyses included genetically inferred Japanese-ancestry participants with WGS; individuals represented in overlapping Japanese AD datasets were removed to limit duplication in meta-analysis.

Illumina WGS reads were aligned to GRCh37, followed by variant calling and joint genotyping with DRAGEN; no imputation was performed. Sample filters addressed sequencing quality, contamination, sex concordance, duplication, heterozygosity and missingness. Genetic PCs were calculated jointly with 1000 Genomes reference samples, retaining participants genetically similar to Japanese or East Asian reference populations. Variants were filtered for sequencing and genotype quality, read depth, call rate, MAF and HWE; X-chromosome HWE testing was restricted to females. Association analyses followed the shared procedures, additionally adjusting for age and sequencing center or batch. Pooled models also included sex. G×S models included genotype and sex main effects and their interaction. Pseudoautosomal and non-pseudoautosomal regions were analyzed separately, with hemizygous male coding and sex-specific X-chromosome quality control.

**National Biobank of Korea**

NBK is operated by the Korea National Institute of Health under the Korea Disease Control and Prevention Agency. AD participants were recruited through the multicenter Biobank Innovations for Chronic Cerebrovascular Disease with Alzheimer's Disease Study (BICWALZS).[24] Cases met criteria for probable AD dementia based on neurological and neuropsychological assessment, including National Institute on Aging–Alzheimer's Association criteria. Structural MRI, amyloid PET or other biomarkers supported diagnoses when available. Controls had no objective cognitive impairment and did not meet criteria for MCI, dementia or other major neurodegenerative disease. Exclusions included other dementia subtypes, major psychiatric disorders, diagnostically confounding conditions, uncertain phenotype status and missing age, sex or covariates. Analyses were restricted to genetically inferred Korean-ancestry participants with WGS, with overlapping Korean AD samples removed to limit duplication.

WGS was performed on Illumina NovaSeq 6000 instruments at an average depth of 30× using 150-bp paired-end reads. Reads were aligned to GRCh37 using BWA-MEM, followed by duplicate marking, base-quality recalibration, variant calling and joint genotyping using DRAGEN. No genotype imputation was performed. Sample filters addressed sequencing depth, contamination, call rate, heterozygosity, sex concordance, duplication and unresolved relatedness. PCs were calculated using common LD-pruned autosomal variants and 1000 Genomes references, retaining participants genetically similar to Korean or East Asian populations. Variant filters included pipeline quality status, genotype depth and quality, allelic balance, call rate ≥98%, MAF ≥0.01 and HWE $P \geq 1 \times 10^{-6}$. X-chromosome HWE testing was restricted to females. Association testing followed the shared procedures.

**Supplementary Methods 2**

**STAGE-AD Transformer: input features, architecture and training**

**Overview**

STAGE-AD is a sex-aware, cross-tissue Transformer for post-GWAS prioritization of AD variants, candidate target genes, tissues and cell types. The harmonized feature space comprised 1,008,420 variants, 19,839 candidate genes, 53 tissue or regulatory compartments and 17 cell types. The model integrated sex-combined, female, male and G×S association statistics with LD, fine-mapping, molecular quantitative trait loci (QTLs), epigenomic annotations and single-cell features. An encoder-only architecture[25] combined modality-specific projections, graph-aware self-attention, sex-conditioned feature modulation and cross-attention across tissue and cellular contexts. Outputs ranked candidate regulatory hypotheses; they did not generate GWAS *P* values or replace formal association testing.

**Input harmonization and sequence construction**

Datasets were harmonized to GRCh38 using chromosome, position, reference allele and alternative allele. Variants with unresolved allele mismatches, ambiguous strand orientation or insufficient quality information were excluded. Each sequence represented an LD-defined locus or fine-mapping interval and contained a learnable [CLS] token, one sex-context token and up to 128 variant, 56 candidate-gene, 53 tissue or regulatory-context and 17 cell-type tokens, giving a maximum length of 256. When the variant cap was exceeded, variants were retained according to fine-mapping probability and association evidence. Candidate genes were retained on the basis of molecular-QTL, enhancer–gene, chromatin-contact and proximity evidence.

Continuous transformations were fitted to the training partition and applied unchanged to validation and test data. Highly skewed variables were transformed, extreme values were bounded, and features were standardized within domains. Missing continuous measurements were assigned a neutral standardized value with a binary missingness indicator; missing categorical annotations received an unknown category. Categorical variables used learnable embeddings. Modality-specific projections mapped inputs to a shared 256-dimensional space, supplemented by token-type, relative genomic-position, source and missingness embeddings. Dataset, ancestry, assay, tissue, cell type, genome build, sample type and processing metadata were retained. Features derived from the same source were grouped to distinguish repeated annotations from independent evidence.

**Genetic association and genomic-context features**

Variant-level association features included effect estimates, standard errors, *z* scores, bounded *P*-value transformations, allele frequencies, effective sample sizes, imputation quality and effect directions across cohorts for sex-combined, female, male and G×S analyses. Additional features represented female–male effect differences, cross-sex direction concordance, between-cohort heterogeneity and sex-stratified replication indicators. *APOE*-stratified and *APOE*-by-sex results were retained separately. Phenotype and analysis-context indicators distinguished clinically diagnosed, biomarker-supported and proxy-defined AD or dementia, as well as clinical-only, proxy-excluded, ancestry-specific and leave-one-cohort-out analyses.

Population-genetic features included ancestry-specific MAF, local variant density, distance from the lead variant, LD-block membership and ancestry-matched LD measures derived from reference panels or cohort genotypes. Ancestry groups were processed separately before integration. Genomic annotations included chromosome, position, variant class, distance to the nearest transcription start site, transcript context and predicted consequence based on GENCODE. Coding, splice-related, untranslated-region, promoter, intronic, intergenic and regulatory annotations were represented alongside protein-altering consequences, predicted loss of function and deleteriousness scores.

Fine-mapping features included conditional-signal and credible-set membership, posterior inclusion probability, credible-set size and the number of distinguishable signals per locus. Ancestry-matched Bayesian estimates were prioritized. Outputs from different fine-mapping methods were retained as method-specific features and consensus indicators rather than averaged.

**Molecular-QTL and regulatory features**

QTL features were compiled from GTEx v8, BrainMeta[26], PsychENCODE[27], AMP-AD, ROSMAP[28], eQTLGen[29] and complementary brain and immune resources. Variant–gene–tissue annotations included eQTL and sQTL effect estimates, association strength, direction, tissue identity and multiple-testing status. Brain regions were retained separately rather than collapsed into a single brain-wide score. Protein, methylation, chromatin-accessibility and histone-modification QTLs were included when supplied by the source datasets. Colocalization probabilities[30] were retained as higher-level regulatory features only when generated without held-out evaluation labels.

Epigenomic features from ENCODE[31], Roadmap Epigenomics[32], PsychENCODE, AMP-AD and single-cell resources included ATAC-seq or DNase-seq accessibility, promoter and enhancer histone marks, chromatin states, transcription-factor binding, DNA methylation and predicted enhancer activity. Annotations retained their tissue, cell type and experimental source and were represented as overlap indicators or quantitative activity scores. Broadly shared annotations were distinguished from tissue- or cell-state-selective elements.

Variant–gene links combined proximity, promoter overlap, enhancer–gene predictions, eQTLs, sQTLs, pQTL, Hi-C, promoter-capture Hi-C and activity-by-contact evidence. Each link retained its mapping method, genomic distance, regulatory direction, biological context and supporting sources. Proximity-only links were distinguished from molecular or chromatin-supported links. Gene features included biotype, transcript structure, expression, sex bias, cell specificity and prior AD-pathway annotations. Candidate genes were not restricted to the nearest gene.

**Tissue, cell-state and sex-context features**

Tissue tokens represented anatomical brain regions, blood and immune compartments, vascular tissues, liver, adipose tissue and other metabolic contexts. Features included expression specificity, QTL activity, chromatin accessibility, enhancer activity, regulatory-element density and the number of supported variant–gene links. Tissue labels were mapped to a controlled ontology. Cellular annotations were derived from single-cell or single-nucleus transcriptomic and accessibility datasets, including multi-region AD atlases, ROSMAP and SEA-AD.[33] Classes included excitatory and inhibitory neurons, microglia, astrocytes, oligodendrocytes, oligodendrocyte precursor cells, endothelial cells, pericytes and peripheral immune populations. Harmonized states represented homeostatic and disease-associated programmes, including lipid-responsive and inflammatory microglia, reactive astrocytes, stressed oligodendrocytes and vascular dysfunction.

Sex-context features included sex-biased expression, differential accessibility, sex-specific molecular-QTL effects, G×S evidence and source-supported hormone-responsive annotations. Estrogen response, menopause-associated metabolic changes, androgen signalling, immune regulation and lipid metabolism were encoded as contextual annotations, not as measurements of circulating hormones. Sex information entered through both a conditioning token and quantitative features. X-chromosome annotations distinguished pseudoautosomal status, male hemizygosity, sex-specific allele frequencies, dosage-compensation assumptions and X-inactivation categories, including escape and variable escape. Available X-chromosome QTL, accessibility and expression features were retained together with indicators of sensitivity to alternative coding assumptions.

**Encoder and sex-conditioning**

The encoder comprised four pre-normalized Transformer layers, each with eight attention heads, a 1,024-dimensional feed-forward network and residual connections. Hidden and embedding dimensions were 256; encoder and embedding dropout were 0.10 and 0.05, respectively. Scaled dot-product attention was defined as

$$\mathrm{Attention}(Q, K, V) = \mathrm{softmax}\left(\frac{QK^{\top}}{\sqrt{d_k}} + M\right)V,$$

where Q, K and V denote query, key and value matrices, $d_k$ is the key dimension, and M combines padding, invalid-edge and graph-derived masks. Edge-type and edge-weight biases encoded prespecified relationships among variants, regulatory elements, genes, tissues and cells. Attention heads were not assigned fixed biological functions.

The sex-context token represented female, male, G×S-interaction or sex-agnostic context. Feature-wise affine modulation[34] generated a conditioned representation,

$$\tilde{h}_i^{(s)} = \gamma_s \odot h_i + \beta_s,$$

which was combined with the shared representation through a sigmoid gate:

$$h_i^{*} = g_i \odot \tilde{h}_i^{(s)} + (1 - g_i) \odot h_i.$$

Contextualized variant tokens then queried tissue and cell-type tokens through cross-attention. Tissue- and cell-aware outputs were fused with the variant and sex-conditioned representations through gated residual connections.

**Prediction heads and learning objectives**

The final [CLS] representation supported binary AD prioritization, five-class sex-context classification (shared, female-biased, male-biased, G×S-interaction and background), candidate-gene, tissue and cell-type ranking, and multilabel regulatory-module prediction. Outputs were interpreted as relative priorities within the candidate set rather than causal evidence.

Masked regulatory pretraining corrupted 15% of observed features or complete tokens using a 70:15:15 mask–random–unchanged scheme. Continuous and categorical inputs were reconstructed using squared-error and cross-entropy losses, respectively. Supervised fine-tuning combined focal modulation ($\gamma = 2.0$)[35] for sparse classification labels, a LambdaRank-compatible gene-ranking objective, listwise tissue and cell-type losses, supervised contrastive learning and Brier-loss calibration. Positive–unlabelled sensitivity analyses used a positive-to-unlabelled sampling ratio of 1:4. The combined objective was

$$\mathcal{L}_{\mathrm{total}} = \sum_t \lambda_t \mathcal{L}_t + \lambda_{\mathrm{mask}}\mathcal{L}_{\mathrm{mask}} + \lambda_{\mathrm{cal}}\mathcal{L}_{\mathrm{Brier}} + \lambda_{\mathrm{con}}\mathcal{L}_{\mathrm{contrastive}}.$$

Task weights were 1.0 for AD prioritization, 1.5 for sex-context classification, 1.0 for gene ranking, 0.4 each for tissue and cell-type ranking, and 0.5 for regulatory-module prediction. Weights for masked reconstruction, calibration and contrastive learning were 0.2, 0.1 and 0.1, respectively. Tasks without labels for an instance were omitted from its normalized loss.

**Data partitioning and optimization**

Chromosome-stratified LD blocks were assigned to training, validation and test partitions in proportions of 70%, 15% and 15%, retaining variants and linked regulatory entities from each LD-defined locus in the same partition. Additional locus, chromosome, connected-component, known-AD-locus, *APOE*-region and chromosome 19 hold-outs assessed generalization to unseen genomic contexts. Chromosome X was retained and evaluated separately, including sensitivity to alternative genotype coding. A synthetic ten-million-variant dataset was used exclusively for software stress testing and input–output validation; synthetic labels and scores were not used as biological results.

Parameters were optimized with AdamW[36] using a peak learning rate of $2 \times 10^{-4}$, minimum learning rate of $1 \times 10^{-6}$, weight decay of 0.01, linear warm-up over 5% of optimization steps and cosine decay. Microbatches of 128 loci and four gradient-accumulation steps gave an effective batch size of 512. Gradient norms were clipped at 1.0, and training used mixed precision. Pretraining and fine-tuning were limited to 50 epochs, with early stopping after seven epochs without improvement in validation AUPRC. Five runs used seeds 2026–2030. Probabilities were calibrated by temperature scaling on validation data.

**Task-specific evaluation and model attribution**

STAGE-AD was compared with 13 prespecified statistical, machine-learning and deep-learning baselines using the same loci, labels, preprocessing and partitions. Variant prioritization was evaluated using AUROC, AUPRC, calibration and precision or recall among the top 100, top 500 and top 1% of predictions. Sex-context classification was evaluated using per-class AUPRC, macro-F1, weighted-F1, balanced accuracy and confusion matrices. Candidate-gene, tissue and cell-type ranking was evaluated using top-k accuracy, mean reciprocal rank and normalized discounted cumulative gain. Pairwise performance differences were estimated from 1,000 bootstrap resamples of held-out loci.

Attribution analyses used integrated gradients[37] and SHAP[38], supplemented by sex-token exchange, 1,000 sex-label permutations, variant and modality masking, and leave-one-tissue- or cell-type-out analyses. Attribution and attention scores were treated as model-dependent, hypothesis-generating measures and required independent statistical-genetic or functional support for biological interpretation. STAGE-AD was implemented in Python with PyTorch. Each run archived the software environment, configuration, data hashes, split manifest, random seed, training log and checkpoint identifier.

**Supplementary Methods 3**

**NPAD multi-omics profiling and neuropathological analyses**

**Cohort and neuropathological assessment**

The Neurodegenerative Pathology and Alzheimer's Disease (NPAD) cohort was established as an independent postmortem brain cohort to investigate sex-related genetic regulatory pathways in AD. The cohort comprised 579 donors, including 312 neuropathologically confirmed AD cases and 267 non-AD controls. Participants underwent clinical assessment before death and neuropathological examination after brain donation. Demographic characteristics, clinical diagnosis, *APOE* genotype, cognitive status and pathological measurements are summarized in the online tables. Informed consent was obtained from each participant, and the NPAD project were each approved by an Institutional Review Board of Jinan University Medical Center (protocol number: 2022031247; approval date: 2022-08-02). The participants also signed an Anatomic Gift Act, and a repository consent to allow their data to be repurposed.

Brain tissues were collected following standardized procedures for postmortem interval (PMI) and tissue preservation. Dorsolateral prefrontal cortex, middle temporal gyrus and hippocampal samples were analyzed (https://www.dropbox.com/scl/fo/mledvqrc3ayt750wwmqe9/ALfaEPlPJSm1GCx-OSYPIlY?rlkey=smsl59g1rtqxekyyl0pcf3xis&st=zfzecdu2&dl=0). Assessments included amyloid-β plaque burden, tau neurofibrillary pathology, Braak stage[39], CERAD score[40], neuritic plaque density, cerebral amyloid angiopathy and neurodegeneration-related measurements. Quantitative traits were normalized within brain region before analysis.

**Genotyping and genetic quality control**

DNA from frozen brain tissue or peripheral blood was genotyped using the Illumina Global Screening Array, covering approximately 700,000 variants. Sample exclusions addressed genotype missingness, abnormal heterozygosity, sex discordance, relatedness and ancestry outliers. Variants with call rate <98%, MAF <0.01, HWE $P < 1 \times 10^{-6}$ or ambiguous strand orientation were excluded. Genotypes were phased with Eagle v2.4 and imputed against TOPMed. Imputed variants required INFO >0.8 and allele-frequency concordance with reference populations. Variants were harmonized to GRCh38 and the discovery GWAS effect alleles. Genetic ancestry was assessed using LD-pruned variants, and the first ten ancestry PCs were retained for downstream adjustment.

**Bulk RNA sequencing**

Total RNA was extracted from frozen brain tissue using silica membrane-based purification and assessed on an Agilent Bioanalyzer. Samples with an RNA integrity number (RIN) <6 were excluded. Approximately 1 µg RNA was used for Illumina TruSeq Stranded mRNA library preparation. Libraries were sequenced on a NovaSeq 6000 with 150-bp paired-end reads, generating approximately 80 million paired-end reads per sample. Reads underwent FastQC assessment and adapter trimming, followed by STAR alignment to GRCh38 and featureCounts quantification using GENCODE v43. Expression data were variance-stabilized, and technical variation was assessed by PCA. Differential expression was analyzed using DESeq2[41] with adjustment for age, sex, *APOE* ε4 status, genetic ancestry, sequencing batch, RIN, PMI and brain region.

**Single-nucleus RNA sequencing**

Nuclei were isolated from frozen tissue using a sucrose-gradient protocol. Approximately 10,000–20,000 nuclei per sample were loaded onto the 10x Genomics Chromium platform, and libraries were prepared using Single Cell 3′ Gene Expression v3.1 chemistry and sequenced on a NovaSeq 6000. Cell Ranger v7.0 aligned reads to GRCh38 and generated gene-by-nucleus count matrices. Nuclei with fewer than 500 detected genes, mitochondrial transcript proportions >10%, abnormal UMI counts or predicted doublets were excluded, leaving approximately 1.2 million nuclei. PCA and UMAP were used for dimensionality reduction and visualization, and Leiden clustering defined cellular populations. Cell identities were assigned using marker genes and reference mapping to human brain atlases. Populations included excitatory and inhibitory neurons, microglia, astrocytes, oligodendrocytes, oligodendrocyte precursor cells, endothelial cells and pericytes. Previously reported signatures quantified microglial lipid responses, astrocyte inflammation, vascular dysfunction and oligodendrocyte stress.

**Chromatin accessibility and H3K27ac profiling**

ATAC-seq[42] was performed on nuclei from matched brain samples using Tn5-mediated fragmentation and adapter insertion, followed by limited-cycle PCR and paired-end NovaSeq 6000 sequencing. Reads were aligned to GRCh38 with BWA-MEM; mitochondrial, duplicate and low-quality reads were removed before MACS2 peak calling. Peaks were annotated using ENCODE, GeneHancer and brain-specific regulatory resources. Differential accessibility was evaluated using a model including genotype, sex, their interaction and covariates:

$$Accessibility \sim Genotype + Sex + Genotype \times Sex + Covariates$$

H3K27ac CUT&Tag[43] was performed using approximately 50,000 nuclei per sample, anti-H3K27ac antibodies and pA-Tn5-mediated tagmentation. Libraries were sequenced on an Illumina NovaSeq platform and aligned to GRCh38. SEACR was used to identify enriched regions, and active enhancer annotations were defined by overlap with H3K27ac peaks and brain-specific enhancer resources. Enhancer–gene relationships were inferred from chromatin-interaction datasets, enhancer–gene databases and eQTL evidence.

**Integration of regulatory evidence**

Genomic, epigenomic, transcriptomic, cellular, proteomic and pathological information was integrated to evaluate variant–regulatory-element–gene–cell-state–pathology evidence paths. Fine-mapped AD variants were linked to regulatory elements through accessibility and enhancer annotations. Variant–gene relationships were evaluated using eQTL, sQTL, chromatin-interaction and colocalization evidence. Gene–cell-state links were assessed using single-nucleus expression signatures and module scores, and module–pathology relationships were tested by regression and mediation analyses. Biological interpretation was restricted to paths with directionally concordant evidence from independently derived sources.

**Pathology association and sex-interaction analyses**

Genetic module scores were related to continuous and binary pathological traits using linear and logistic regression, respectively. The primary linear model included module score, sex, their interaction and covariates:

$$Y_i = \beta_0 + \beta_1 S_i + \beta_2 Sex_i + \beta_3 (S_i \times Sex_i) + \sum_k \gamma_k C_{ik} + \varepsilon_i.$$

Here, $S_i$ denotes the participant-level module score and $C_{ik}$ the covariates. Covariates comprised age at death, *APOE* ε4 status, ancestry PCs, brain region, PMI, RIN and sequencing batch; sex was included explicitly as a main effect. The module-score-by-sex coefficient assessed heterogeneity of the association between sexes. Repeated measurements across brain regions and technical batches were additionally analyzed with mixed-effects models containing donor and batch random intercepts. Molecular association tests were adjusted using the Benjamini–Hochberg false discovery rate procedure.

**Mendelian randomization and mediation**

Two-sample Mendelian randomization (MR) was used to assess potential directional relationships between genetically predicted module activity and pathological phenotypes. Instruments were selected using the prespecified association threshold and LD $r^2 < 0.01$; instruments with $F < 10$ were excluded. Inverse-variance weighting[44] was the primary estimator. Sensitivity analyses comprised weighted-median and MR-Egger estimation[45], MR-PRESSO[46] outlier assessment, Cochran's Q, the MR-Egger intercept, Steiger directionality testing and leave-one-out analyses.

Statistical mediation analyses evaluated whether intermediate cellular states accounted for associations between genetic modules and pathology. Indirect effects were estimated using the product of coefficients, with confidence intervals derived from 5,000 bootstrap iterations. These analyses were interpreted as evidence of statistical mediation compatible with the proposed pathways, rather than proof of biological causality.
